# Gut microbial structural variation improves disease discrimination and reveals a strain-level regulatory mechanism in rheumatoid arthritis

**DOI:** 10.64898/2026.07.23.26358822

**Authors:** Yang Ou, Shengyan Zhao, Jiachun Weng, Yichen Long, Huamei Li, Yue Hou, Qing Xiong, Sha Liu, Zhikang Wang, Yuhui Xu, Hui Pan, Huayong Zhang, Lingyun Sun

## Abstract

Gut microbial structural variation captures strain-level genomic diversity beyond species abundance, yet its contribution to rheumatoid arthritis (RA) remains unclear. Integrating gut metagenomic datasets from four independent cohorts (n = 491), we identified reproducible RA-associated structural variants (SVs), most of which occurred in species without differential abundance. Incorporating SVs into machine learning models consistently improved disease discrimination across independent validation cohorts. Functional analyses identified a core deletion SV in *Agathobacter rectalis* that removes an XRE-family transcriptional regulator. Motif discovery and 3D structural modeling demonstrated sequence-specific binding of the regulator to the promoter of a short-chain fatty acid biosynthetic gene, supporting a strain-level regulatory mechanism. Together, these findings establish microbial structural variation as a complementary functional layer beyond taxonomy for RA discrimination and mechanistic interpretation.

## Introduction

Rheumatoid arthritis (RA) is a chronic systemic autoimmune disease characterized by persistent synovial inflammation and progressive joint destruction^1^. Its pathogenesis involves complex interactions between genetic susceptibility and environmental factors^2^. Increasing evidence has implicated the gut microbiome in RA susceptibility and immune dysregulation^3^. Alterations in intestinal microbial composition and function have been associated with impaired immune tolerance, systemic inflammation, and autoantibody-associated immune responses, including anti-citrullinated protein antibodies (ACPAs), which may precede clinical disease onset ^4,5^.

Although microbiome-targeted interventions such as fecal microbiota transplantation (FMT)^6,7^ and probiotic supplementation^8,9^ aim to restore microbial homeostasis, their variable efficacy highlights host–microbiome complexity and heterogeneity. A major limitation of current microbiome studies is that conventional taxonomic profiling often fails to capture functional variation within bacterial species. Thus, identifying high-resolution metagenomic biomarkers beyond species-level abundance remains an important challenge.

Recent advances in metagenomics have revealed that microbial structural variants (SVs), including deletion structural variants (dSVs) and variable structural variants (vSVs), represent an important source of strain-level functional diversity^10^. These regions can harbor genes involved in microbial adaptation and host–microbe interactions, providing functional resolution beyond taxonomy. Previous studies have demonstrated that microbial SVs are associated with host metabolic traits and circulating metabolomic profiles, suggesting their potential to capture biologically relevant microbiome variation that is not reflected by taxonomic composition alone^10,11^.

Despite the established association between gut microbial dysbiosis and RA, the contribution of microbial SVs to RA remains largely unexplored. In particular, it remains unknown whether strain-level genomic variation provides additional discriminatory information beyond species-level taxonomy and whether RA-associated SVs are linked to specific regulatory mechanisms influencing microbial functions. Addressing these questions may reveal an additional functional layer connecting microbial genomic variation with immune-relevant processes in RA.

To address these gaps, we performed a cross-cohort meta-analysis integrating gut metagenomic datasets from four independent RA cohorts. Beyond conventional taxonomic profiling, we systematically characterized microbial SV landscapes and evaluated their contribution to RA discrimination beyond species-level abundance. Furthermore, we identified a key SV-associated transcriptional regulator and investigated its potential regulatory mechanism through motif analysis and experimental DNA-binding validation. Together, our findings establish microbial structural variation as an additional functional layer of the gut microbiome associated with RA and highlight its potential for disease discrimination and functional interpretation.

## Results

### Microbial Diversity and Clinical Profiles Across Cohorts

Clinical characteristics were largely comparable across the four cohorts (Table S1). RA patients were predominantly female (67.5%–82.9%) with median ages ranging from 51 to 60 years. In Cohort D, untreated RA patients exhibited higher inflammatory activity, reflected by elevated erythrocyte sedimentation rate (ESR) and C-reactive protein (CRP) levels compared with treated patients and healthy controls. High ACPA seropositivity was consistently observed across RA groups, particularly among untreated individuals.

Microbial alpha diversity was generally comparable between RA and control groups, with reduced observed richness detected only in Cohort A (Table S2, Figure S1). Other diversity metrics, including Shannon diversity, Simpson index, and Pielou’s evenness, showed no significant differences across groups. Beta diversity analysis based on Bray–Curtis dissimilarity revealed cohort-dependent differences in microbial community composition. Significant compositional shifts were observed in Cohorts A and C, whereas no significant differences were detected in Cohorts B and D (Table S3, Figures S1–S4). These findings indicate variability in community-level microbial profiles across RA cohorts.

### Landscape of Gut Microbial Structural Variants Across Cohorts

To investigate genomic variation beyond species-level composition, we characterized microbial structural variants (SVs) across the four cohorts. A total of 4,105 SVs were identified, including 2,860 variable SVs (vSVs) and 1,245 deletion SVs (dSVs) (Figure 2A, B). SVs displayed strong species specificity, with *Blautia wexlerae*, *Roseburia intestinalis*, and *Agathobacter rectalis* harboring the highest numbers of detected SVs.

**Figure 2.**
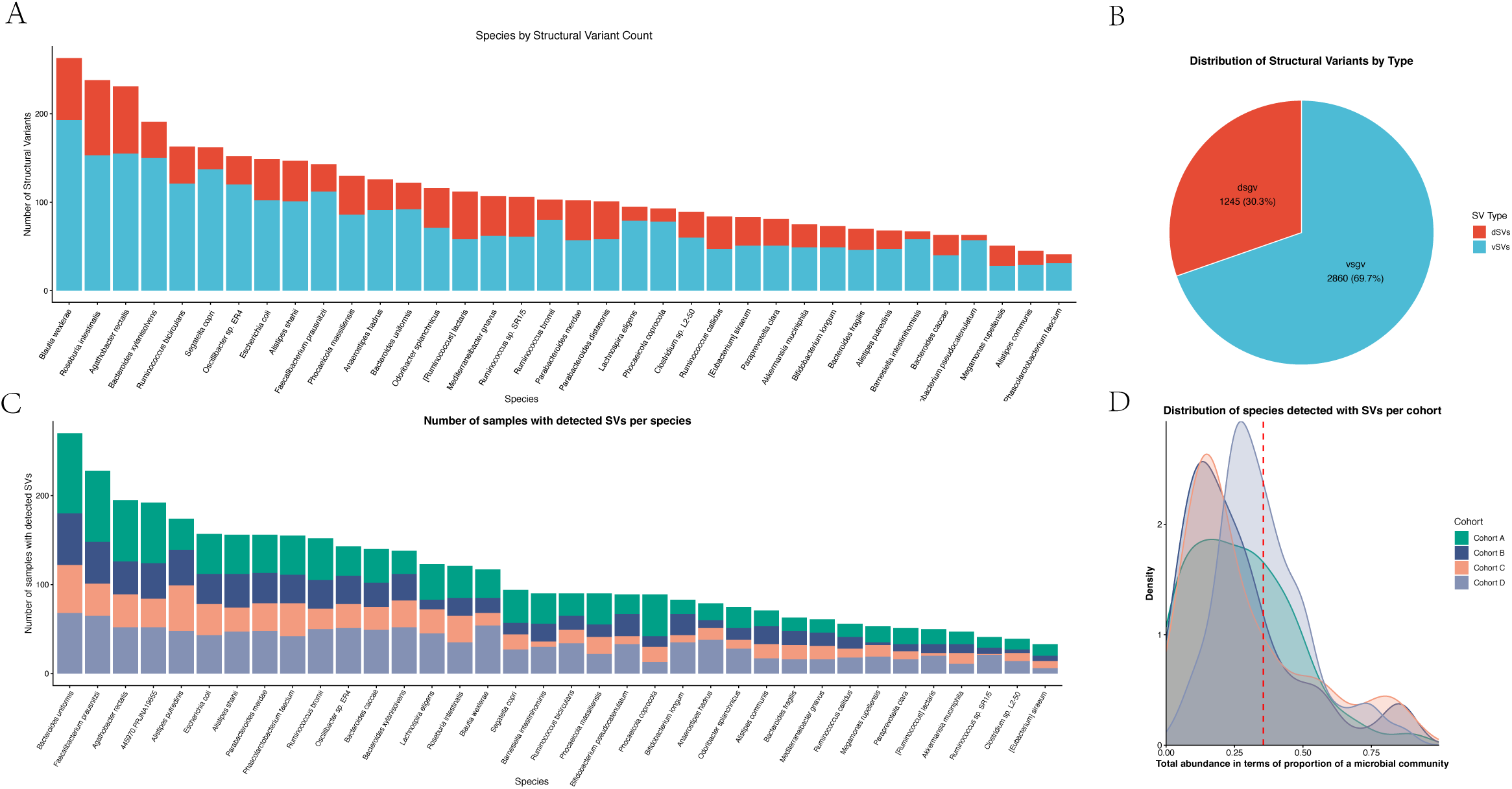
Landscape, distribution, and cohort-level prevalence of gut microbial structural variants (SVs). (A) Distribution of detected SV counts across bacterial species, color-coded by SV classification. (B) Proportional distribution of identified SV types, highlighting deletion SVs (dSVs, 30.3%) and variable SVs (vSVs, 69.7%). (C) Number of human stool samples harboring detected SVs per species, stratified across independent study cohorts. (D) Relative abundance density distributions of bacterial species carrying detected SVs within microbial communities across cohorts.

The distribution of SV-harboring species varied across cohorts, with *Bacteroides uniformis* and *Faecalibacterium prausnitzii* identified in the largest number of SV-detected samples (Figure 2C). In addition, species detected with SVs constituted a major fraction of the total microbial community (median cumulative abundance ∼35%), exhibiting remarkably consistent density distributions across the four cohorts (Figure 2D).

### Meta-analysis Reveals Consensus Microbial Taxonomic and Structural Signatures

To identify robust RA-associated microbial features across populations, we performed a meta-analysis integrating four independent cohorts. This analysis identified 49 species with consistent differential abundance (Table S4, *P_meta_* < 0.05, *P_heterogeneity_* > 0.01) (Figure 3A). At the genomic level, we further identified 34 vSVs and 25 deletion dSVs significantly associated with RA (Table S5, S6 and Figure 3B, C).

**Figure 3.**
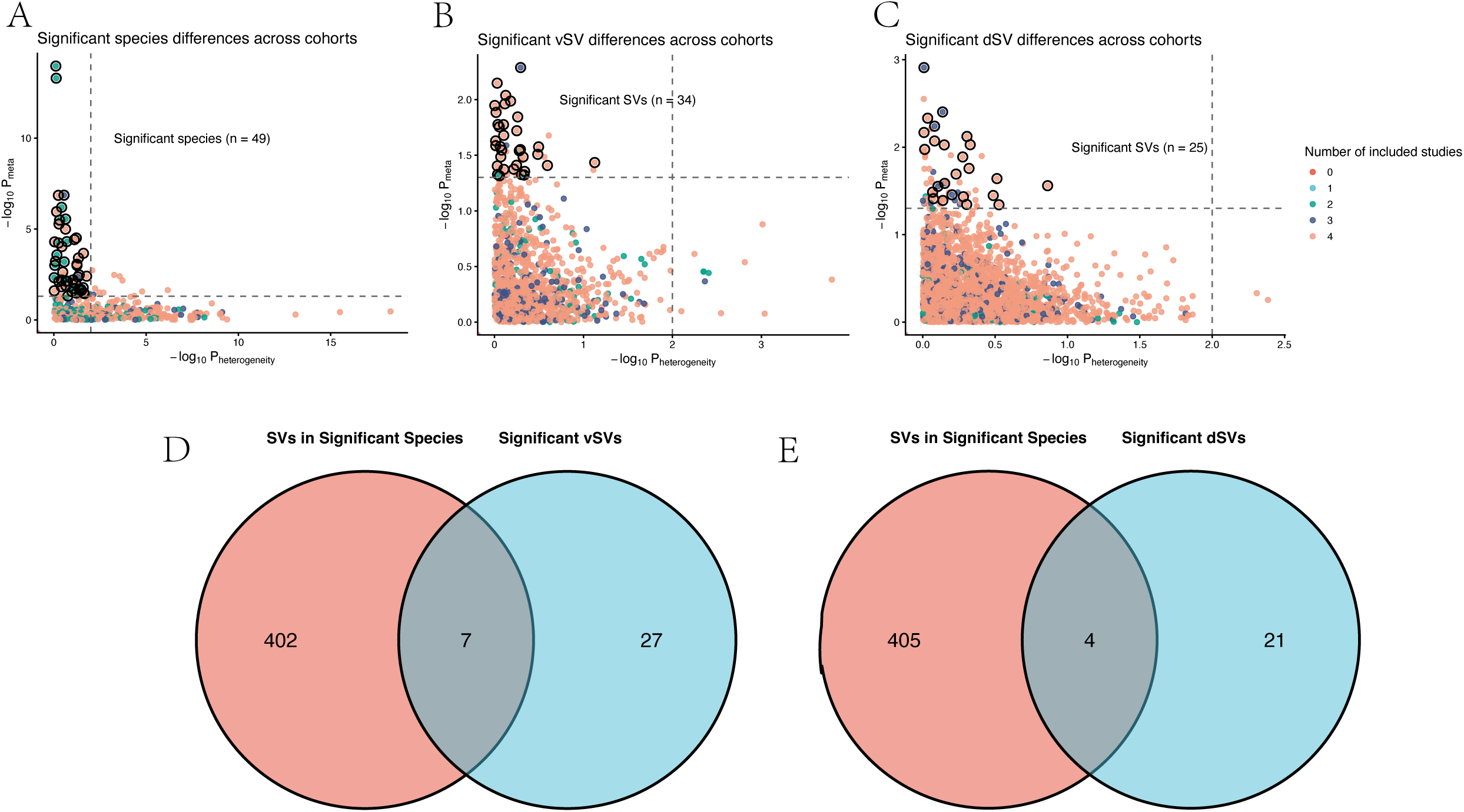
Meta-analytic discovery and overlap of species-level and structural variant-level differences across multi-center cohorts. (A–C) Volcano plots displaying random-effects meta-analysis results for species-level abundance differences (A), variable SVs (vSVs) (B), and deletion SVs (dSVs) (C) across cohorts. Horizontal dashed lines denote significance thresholds (*P_meta_* < 0.05) and vertical dashed lines mark heterogeneity limits (*P_heterogeneity_*). Point sizes indicate the number of included studies. (D, E) Venn diagrams illustrating the overlap between SVs located within significantly altered bacterial species and significantly differentiated vSVs (D) or dSVs (E).

Interestingly, the vast majority of significant SVs did not reside within differentially abundant species, with only 7 vSVs and 4 dSVs overlapping with species-level abundance changes (Figure 3D, E). This decoupling indicates that microbial structural variations provide additional disease-associated information beyond conventional taxonomic profiles.

### Machine Learning Performance and Cross-Cohort Validation of Consistently Prioritized Structural Variants

To evaluate whether microbial SVs improve classification performance for RA while avoiding discovery bias, Cohort A was reserved strictly for candidate SV identification, with model training performed on independent datasets (Cohort B+C for training, Cohort D for external validation). Incorporating SVs markedly enhanced classification performance over species abundance alone across all cohorts (Figure 4A–F, Table S7). Under the λ_1se_ parameterization, the Species+SV model achieved area under the curves (AUCs) of 0.82 in the training cohort (Cohort B+C; vs. 0.71 for Species-only, *P* < 0.01; Figure 4A), 0.62 in the independent test cohort (Cohort D; vs. 0.48, DeLong’s test *P* = 0.04; Figure 4B, Table S7), and 0.78 in the discovery cohort (Cohort A; vs. 0.65, *P* = 0.02; Figure 4C). Under the λ_min_ parameterization, similar gains were observed, with AUCs reaching 0.83 in training (vs. 0.77, *P* < 0.01; Figure 4D), 0.60 in test (vs. 0.45, DeLong’s test *P* < 0.01; Figure 4E, Table S7), and 0.79 in discovery (vs. 0.71, *P* = 0.04; Figure 4F).

**Figure 4.**
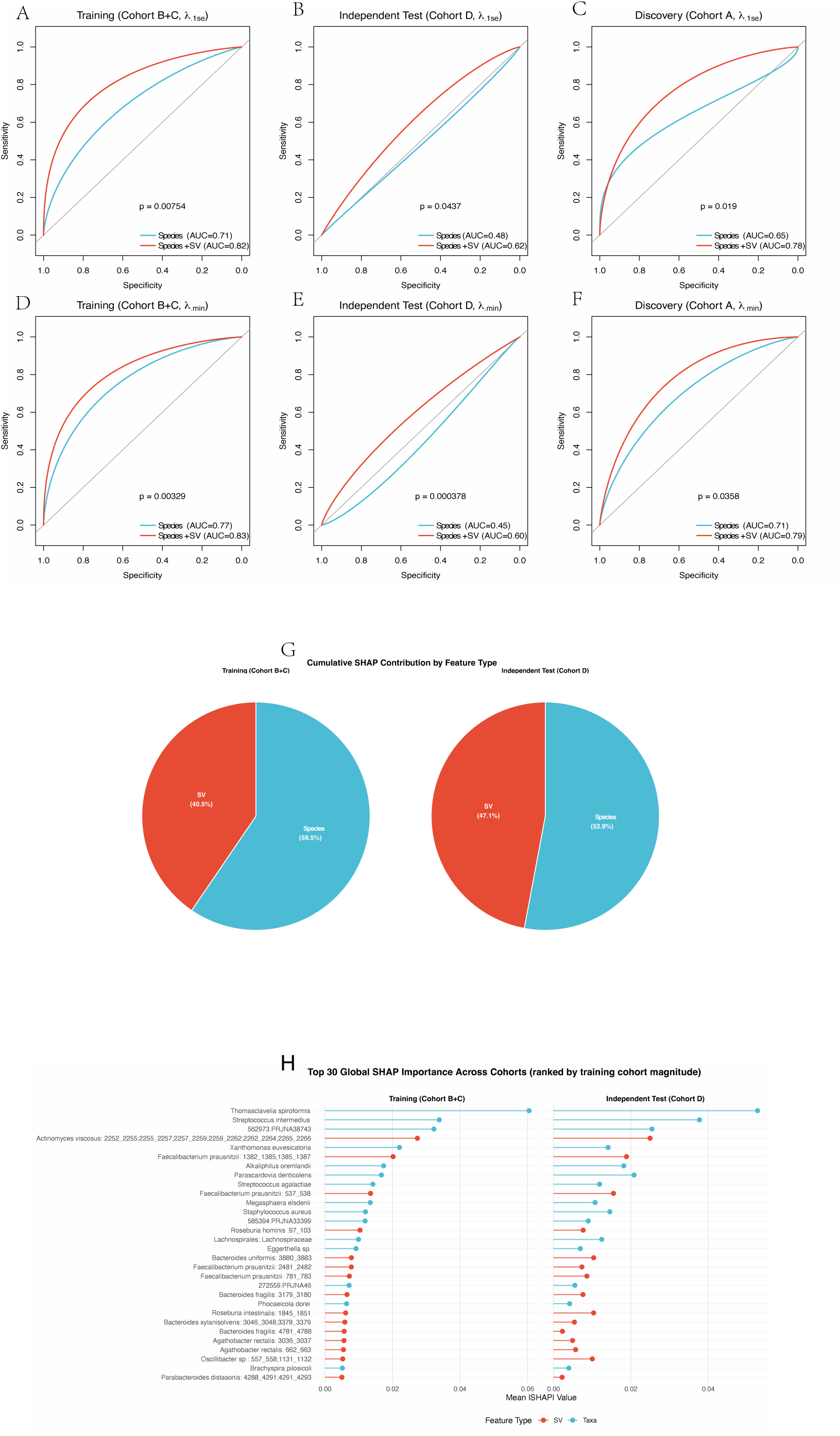
Cross-cohort performance, classification discrimination, and global feature importance of microbial SV machine learning models. (A–F) Receiver operating characteristic (ROC) curves evaluating disease classification performance using species-only models (blue) versus combined species-plus-SV models (red) across Training Cohort B+C (A, D), Independent Test Cohort D (B, E), and Discovery Cohort A (C, F) evaluated under alternate regularization parameters (λ_1se_ and λ_min_). Area Under the Curve (AUC) values and DeLong test P-values are indicated. (G) Cumulative SHAP contribution pie charts comparing the overall predictive weight of SV features versus species features in training and test cohorts. (H) Top 30 global SHAP importance ranking of predictive features across cohorts, ranked by mean absolute SHAP value in the training cohort (B + C).

SHapley Additive exPlanations (SHAP) feature importance analysis in the training cohort confirmed that SV features contributed substantially to model predictions, accounting for 40.50% of the cumulative feature importance (and 47.70% in the test cohort; Figure 4G). Top global features ranked by training magnitude highlighted key specific SVs across cohorts (Figure 4D). Cross-cohort SHAP profile alignment (Figure 5A, B) and quantile-scaled SHAP summary distributions (Figure S5) prioritized robust RA-associated features based on consensus criteria (Table S8), including *Alistipes shahii*: 2252_2255;2255_2257;2257_2259;2259_2262;2262_2264;2265_2266, *Bacteroides uniformis*: 3880_3883, and *Bacteroides fragilis*: 4781_4788 (Spearman r = 0.69 to 0.91, pooled ORs = 1.86 to 4.18; Table S8, Figures S6, 7). Conversely, persistent protective factors such as *Faecalibacterium prausnitzii*: 537_538, *Faecalibacterium prausnitzii*: 781_783, *Agathobacter rectalis*: 662_663, and *Ruminococcus bromii*: 97_103 were identified (Spearman r = -0.67 to -0.89, pooled ORs = 0.34 to 0.53; Table S8, Figures S6, 7). Notably, leave-one-out sensitivity analyses confirmed that two key SVs maintained robust cross-cohort stability (Figures S8, 9). In subgroup analyses, *Agathobacter rectalis*: 662_663 showed significant frequency differences in both osteoarthritis (OA) vs. RA and Untreated vs. Treated RA comparisons (*P* = 0.04 and *P* = 0.03, respectively; Figure 5C, D), whereas *Faecalibacterium prausnitzii*: 781_783 showed no significant differences across these subgroups (*P* = 1.00; Figure S10).

**Figure 5.**
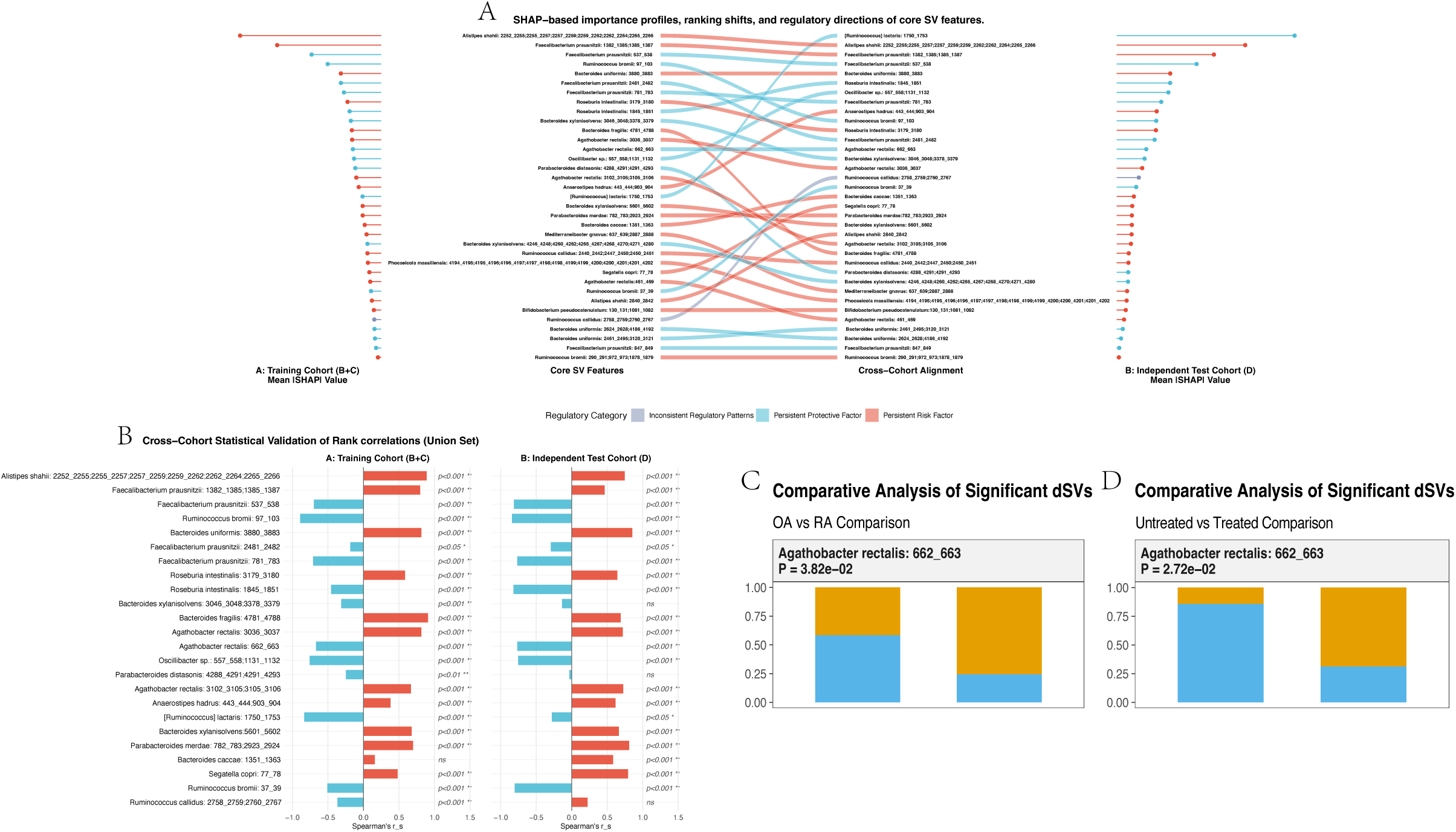
Cross-cohort alignment, ranking shifts, and regulatory consistency of consistently prioritized SV features. (A) Cross-cohort statistical validation and alluvial plot mapping rank shifts and mean |SHAP| importance values of top SV features (union of top 20 ranked features in Training Cohort B+C or Independent Test Cohort D). Features are color-coded by regulatory directionality, highlighting persistent protective factors, persistent risk-associated features, and inconsistent regulatory patterns. (B) Spearman correlation matrices illustrating cross-cohort consistency and alignment of feature importance rankings. (C) Comparative prevalence analysis of representative dSVs (*Agathobacter rectalis*: 662_663) between disease subgroups (OA vs. RA, left; Untreated vs. Treated RA, right).

### Functional Annotation and Structural Characterization of Consistently Prioritized Deletion SVs

To explore the strain-level functional impacts of consistently prioritized deletion SVs, we analyzed their genomic locus structures, conserved functional domains, and locus-specific regulatory potential based on sequence alignments and structural predictions.

For *Faecalibacterium prausnitzii*: 781_783, domain significance analysis revealed that this deletion region encodes a bacterial retron-associated reverse transcriptase element (HMPREF9436_01829) dominated by cd03487: RT_Bac_retron_II (-log_10_ E-value = 54.32) and NF038233: retron_St85_RT (-log_10_ E-value = 36.51) (Figure S11). Detailed site profiling mapped conserved catalytic centers, including putative active sites, NTP-binding sites, and nucleic acid-binding sites (Figure S11). Strain-level phylogenetic mapping illustrated the structural loss of this target gene across specific *Faecalibacterium prausnitzii* strains (Figure S12). Mechanistically, retron-encoded reverse transcriptases synthesize multicopy single-stranded DNA (msDNA) to act as molecular sensors in bacterial antiphage defense systems, triggering abortive infection to halt viral replication upon phage invasion. Because *Faecalibacterium prausnitzii* is a major butyrate-producing commensal involved in maintaining gut mucosal integrity and immune homeostasis, the structural loss of this retron-associated locus in RA may indicate altered strain-level bacterial defense capacity and ecological persistence, with potential consequences for host–microbe interactions.

For *Agathobacter rectalis*: 662_663, domain architecture profiling demonstrated that this SV region uniquely harbors a bacterial transcriptional regulator, *EUR_RS03225*, belonging to the Xenobiotic Response Element family (Figure 6A, B). It is characterized by COG1476: XRE (-log_10_ E-value = 23.56) and smart00530: HTH_XRE (-log_10_ E-value = 13.56), containing sequence-specific DNA-binding sites and a salt bridge (Figure 6A, B). Strain-level phylogenetic mapping illustrated the differential presence/absence of this locus across *Agathobacter rectalis* isolates (Figure 6C). Base-level genomic profiling (Pos: 371814) located the target binding motif upstream of the target gene *EUR_RS01730* (Figure 6D). Hypothesizing that XRE-family proteins commonly mediate negative feedback autorepression through binding to their own promoters^12^, we sought to systematically decipher the consensus DNA-binding preference of EUR_RS03225. We identified homologous proteins of EUR_RS03225 across *Agathobacter rectalis* and related strains using BLASTP, extracted the 500-bp upstream promoter regions of their encoding genes, and performed de novo motif discovery via the MEME suite. This analysis identified a highly conserved consensus DNA-binding motif enriched within these promoter regions (top motif E-value = 7.20E-88) (Figure S13). Notably, the predominant motif exhibited a characteristic palindromic (inverted repeat) structure, consistent with the dimeric DNA-binding mode of XRE-family regulators, supporting the presence of a self-regulatory promoter motif that facilitates tight negative feedback control over *EUR_RS03225* expression.

**Figure 6.**
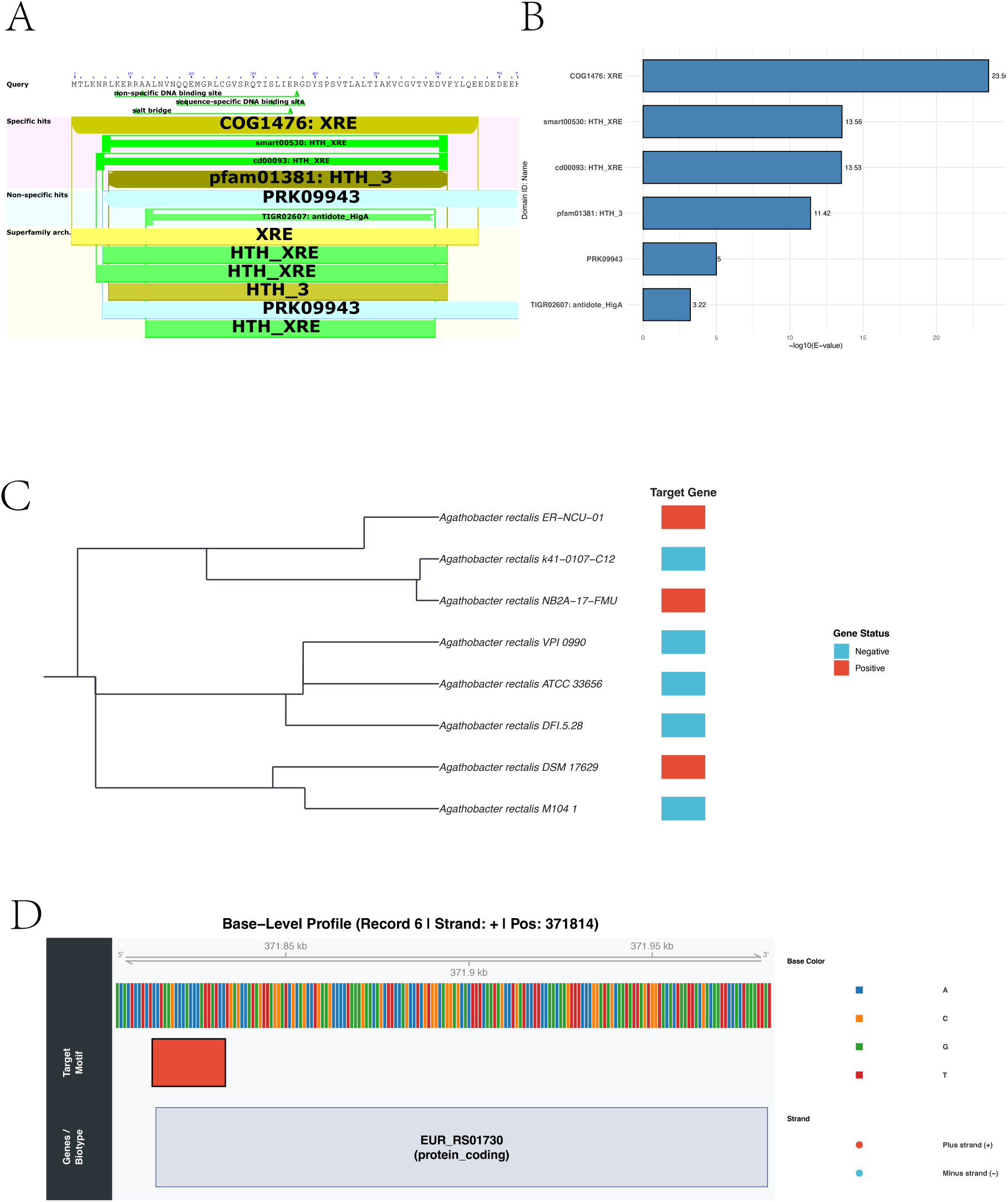
Functional domain annotation, catalytic profiling, and genomic architecture of the SV-associated transcriptional regulator EUR_RS03225. (A) Conserved domain architecture and sequence site profiling of *EUR_RS03225*, highlighting DNA-binding domains, salt bridges, and specific contact sites. (B) Domain significance analysis indicating primary hits dominated by COG1476: XRE (-log_10_ E-value = 23.56) and smart00530: HTH_XRE (-log_10_ E-value = 13.56). (C) Strain-level phylogenetic distribution showing presence (red) or absence (blue) of the target regulator locus across *Agathobacter rectalis* isolates. (D) Base-level genomic profile illustrating the upstream target binding motif located within upstream regulatory region the transcription locus of downstream gene *EUR_RS01730*.

At this motif binding locus, detailed genomic mapping identified a specific downstream gene whose transcription start site is directly overlapped by the binding motif (Figure 6D). Domain annotation of this target locus and adjacent metabolic genes highlighted key acetyl-CoA acetyltransferase elements (TIGR01930: AcCoA-C-Actrans, PRK05790, PaaJ, and thiolase, -log10 E-value = 300.0) (Figure S14). Because thiolase-mediated acetyl-CoA condensation is the critical rate-limiting enzymatic step in SCFA biosynthesis, particularly butyrate production, this genomic configuration suggests a potential regulatory relationship between EUR_RS03225 and genes involved in butyrate-associated metabolism. 3D structural modeling and molecular docking of the SV-associated regulator dimer (EUR_RS03225) with target DNA predicted stable interaction models for wild-type sequences (ipTM = 0.74, pTM = 0.79 for EUR_RS03225_WT; ipTM = 0.72, pTM = 0.77 for EUR_RS01730_WT), whereas sequence mutations reduced predicted binding affinity (ipTM = 0.67 and 0.69, respectively) (Figure S15). Depletion of this regulator cassette could impair microbial butyrate biosynthesis, leading to reduced production of an immunomodulatory metabolite involved in regulatory T cell induction and mucosal barrier function, thereby providing a potential mechanism linking microbial structural variation to immune dysregulation in RA.

## Discussions

In this study, we systematically characterized gut microbial structural variation in rheumatoid arthritis by integrating metagenomic data from four independent cohorts. Beyond conventional species-level profiling, we demonstrated that microbial SVs represent an additional layer of microbiome variation associated with RA and capture disease-related genomic heterogeneity that is not reflected by taxonomic abundance alone. Cross-cohort meta-analysis identified reproducible RA-associated microbial signatures at both species and SV levels, while machine learning analyses showed that incorporation of SV features improved discrimination between RA patients and controls beyond species-level profiles. Furthermore, functional characterization of representative SVs revealed strain-level genomic alterations affecting microbial regulatory and defense-related loci, providing potential biological insights into how microbial genomic variation may contribute to RA-associated microbiome states. Together, these findings highlight microbial structural variation as an underexplored dimension of the gut microbiome with potential relevance for biomarker discovery and functional interpretation in RA.

Previous studies have established an association between gut microbiome alterations and RA, with recurrent observations including the expansion of *Prevotella copri*^13^ and increasing evidence supporting immunomodulatory roles of commensal taxa such as *Faecalibacterium prausnitzii* in regulating inflammatory responses and intestinal immune homeostasis^14^. However, species-level abundance profiling provides only a partial representation of microbial variation, as bacterial species frequently comprise genetically diverse populations with substantial strain-level heterogeneity^15^. Strains belonging to the same species can differ extensively in their accessory genomes, including gene gain and loss events, resulting in functional divergence in metabolic capacity, ecological adaptation, and host–microbe interactions^10^. Therefore, changes in species abundance may capture ecological shifts within microbial communities while overlooking genomic heterogeneity among closely related bacterial populations. Microbial SVs, which represent strain-level differences in genome organization and gene content, provide an additional genomic layer capable of capturing functional diversity beyond conventional taxonomic profiling^10^. By resolving microbial variation at higher genomic resolution, SV-based approaches may reveal disease-associated microbial features that remain undetected when analyses are restricted to species-level abundance^10^.

Beyond characterizing disease-associated microbial alterations, metagenomic studies have increasingly explored the potential of microbiome-derived features as biomarkers for complex diseases. Previous studies have demonstrated that gut microbial signatures can discriminate individuals with metabolic disorders, inflammatory diseases, and other clinical phenotypes, highlighting the potential of microbiome-based classifiers for disease stratification^16,17^. However, the translational application of microbiome biomarkers remains challenging due to substantial inter-individual variability, population heterogeneity, and limited reproducibility across independent cohorts^18,19^. Moreover, most existing microbiome classifiers rely primarily on taxonomic abundance profiles, which may overlook functionally relevant genomic differences among strains within the same species. In this study, we found that incorporation of microbial SV features improved classification performance beyond species-level abundance profiles and identified reproducible SV-associated signatures across independent RA cohorts^15^. These findings suggest that microbial genomic variation may complement conventional taxonomic features and provide additional resolution for microbiome-based biomarker development in RA.

Beyond their potential biomarker value, microbial SVs may represent biologically meaningful genomic alterations rather than merely statistical features. Emerging studies have demonstrated that bacterial SVs can capture strain-level functional variation associated with diverse host phenotypes. For example, microbial SVs have been linked to host metabolic traits through strain-specific gene content variation, revealing functional interactions between host genetic background and microbial genomic diversity^20,21^. In addition, disease-associated microbial SV signatures have been identified in children with autism spectrum disorder and were shown to improve disease classification beyond species-level abundance profiles^22^. Furthermore, microbial SVs have been associated with response to immune checkpoint inhibitor therapy, suggesting that strain-level genomic variation may influence host immune-related phenotypes^23^. Together, these findings indicate that microbial SVs represent an emerging functional layer of the microbiome that extends beyond conventional taxonomic composition. However, whether microbial SVs contribute to autoimmune diseases such as RA remains largely unexplored.

In our study, functional annotation of RA-associated SVs revealed alterations affecting diverse microbial functional modules. For example, deletion of a retron-associated reverse transcriptase locus in *Faecalibacterium prausnitzii* suggests that disease-associated SVs may influence bacterial defense-related functions, highlighting the potential functional consequences of strain-level genomic variation^24^. More notably, deletion of the XRE-family transcriptional regulator cassette in *Agathobacter rectalis* provided an example of how microbial SVs may affect bacterial regulatory architecture. XRE-family regulators are sequence-specific DNA-binding proteins involved in transcriptional regulation, and loss of such regulatory elements may alter bacterial gene regulatory networks^25^. In our study, motif analysis, structural modeling, and DNA-binding validation supported the interaction between the XRE-family regulator and a conserved promoter-associated motif, suggesting that this SV-associated deletion may influence regulatory capacity at the strain level. Although the downstream metabolic consequences require further investigation, the overlap of the predicted binding motif with the transcriptional start site of a metabolic gene provides a plausible framework linking microbial genomic variation with altered regulatory potential and host-relevant microbial functions^26^.

Several limitations should be acknowledged. First, although we integrated four independent cohorts and applied a standardized analytical framework, the retrospective nature of publicly available metagenomic datasets limits the ability to establish temporal relationships between microbial SV alterations and RA development. Prospective longitudinal studies will be required to determine whether specific SV signatures precede disease onset or change during disease progression and treatment. Second, while functional analyses and molecular validation of the XRE-family regulator support its sequence-specific DNA-binding activity and the biological relevance of this SV-associated regulatory locus, future studies evaluating its downstream metabolic consequences, including potential effects on butyrate-associated metabolism, will be important to further clarify its functional impact. Finally, larger cohorts integrating microbial genomic features with clinical characteristics, host genetic factors, and immune phenotypes will be important to evaluate the translational potential of SV-based microbiome signatures in RA.

In conclusion, our study expands the current understanding of RA-associated microbiome alterations by demonstrating that microbial structural variation provides information beyond species-level taxonomy. These strain-level genomic features improve characterization of disease-associated microbial states and reveal functional genomic changes that may otherwise remain undetected by conventional abundance-based approaches. By establishing microbial SVs as a complementary layer of microbiome variation, our findings provide a framework for future studies investigating microbial genomic heterogeneity, biomarker development, and host– microbiome interactions in rheumatoid arthritis.

## Methods

### Data Acquisition and Cohort Integration

This study utilized publicly available gut metagenomic sequencing data from four clinical cohorts of RA patients, totaling 491 samples. Cohort A (Cheng et al., 2022) focuses on the comparison between RA patients, osteoarthritis (OA) patients, and healthy controls (HC)**^27^**, Cohorts B and C (He et al., 2022) investigate the differences between RA patients and healthy individuals**^28^**, and Cohort D (Zhang et al., 2015) provides a comprehensive comparison between healthy controls and RA patients, the latter of which includes both disease-modifying antirheumatic drug (DMARD)-treated and untreated individuals**^29^**. Raw metagenomic reads were obtained from the Genome Sequence Archive (GSA) and the European Bioinformatics Institute (EBI) database, while clinical metadata were retrieved from the supplementary materials of the original publications (Table S1). To ensure cross-cohort consistency, all sequence data were reprocessed using a standardized metagenomic workflow based on unified host-read removal, taxonomic profiling, and structural variation identification procedures, with comprehensive baseline clinical data detailed in Figure 1.

### Metagenomic Preprocessing and Quality Control

The raw metagenomic sequencing data were subjected to quality control and host-read depletion before downstream microbial profiling. Adapter sequences and low-quality reads were trimmed using Trimmomatic (v0.39)^30^ with the parameters LEADING:3, TRAILING:3, SLIDINGWINDOW:4:20, and MINLEN:50. Host-derived reads were identified and removed by mapping the processed reads to the human reference genome (GRCh37) using Bowtie2 (v2.3.5.1)^31^ in --very-sensitive mode. The preprocessing workflow was performed using the KneadData (v0.6.1) wrapper^32^, generating host-depleted microbial reads for subsequent taxonomic profiling and structural variation analysis.

### Identification of Microbial Structural Variations

Microbial SVs, including dSVs and vSVs, were identified using SGV-Finder based on the Iterative Coverage-based Read Assignment (ICRA) algorithm to resolve ambiguous read mappings^10^. Microbial genomes from the proGenomes database^33^ were partitioned into 1-kbp bins based on concatenated genome scaffolds for coverage-based SV identification. SV discovery was initially performed in Cohort A (n = 122) using the --min_samp_cutoff parameter set to 12, corresponding to approximately 10% of the cohort size, to identify prevalent structural variants while minimizing rare sample-specific signals. For replication analyses, the SV status of candidate SV regions identified in Cohort A^27^ was quantified in Cohorts B, C^28^, and D**^29^** using SGV-Finder with the --by-orig parameter. These SVs were subsequently evaluated for disease associations through cohort-specific statistical analyses and meta-analysis.

### Taxonomic Quantification and Diversity Analysis

Taxonomic profiles were generated by aligning quality-controlled metagenomic reads to the proGenomes database^33^ using Bowtie2 (default settings)^31^, followed by processing with Samtools^34^ and CoverM^35^ to generate species-level abundance profiles, including raw read counts, covered fraction, mean coverage depth, TPM, and RPKM values. To minimize noise, microbial features present in fewer than five samples within a cohort were excluded from downstream analysis. Raw read counts were used for differential abundance analysis, whereas normalized abundance profiles were used for microbial diversity analyses.

Alpha diversity indices, including Shannon diversity, Simpson diversity, and observed richness, were calculated based on filtered abundance profiles. Beta diversity was assessed using Bray–Curtis dissimilarity calculated from relative abundance data via total sum scaling (TSS) and visualized using Non-metric Multidimensional Scaling (NMDS) and Principal Coordinates Analysis (PCoA), with statistical significance evaluated by PERMANOVA.

### Statistical Analysis and Consensus Meta-Analysis

Differential abundance analysis was performed using the edgeR quasi-likelihood (QL) framework**^36^** based on raw read counts with Trimmed Mean of M-values (TMM) normalization to account for differences in library size across samples.

To identify consistent microbial signatures across cohorts, random-effects meta-analyses were performed using the metafor^37^ and meta^38^ packages. Species-level differential abundance results were integrated using restricted maximum likelihood (REML)-based models. Binary dSV associations were summarized as odds ratios using Mantel–Haenszel (MH) random-effects meta-analysis. Continuous vSVs were analyzed as standardized mean differences (SMDs) and subsequently converted to log odds ratios using the formula 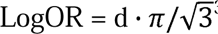^39^. Consensus significance was defined by a three-tier filtering system requiring a pooled meta-analysis *P* ≤ 0.05, a heterogeneity test *P* > 0.01, and replication in at least two independent cohorts with a nominal *P* ≤ 0.2 and a consistent effect direction^23^.

### Machine Learning Model Construction and Feature Interpretation

To evaluate whether microbial structural variants provided additional discriminatory information beyond species-level abundance, machine learning models were constructed using meta-analysis-derived significant microbial features. Cohort A was reserved as the discovery cohort for SV identification, whereas Cohorts B and C were combined as the training dataset and Cohort D was used as an independent validation dataset.

Prior to model construction, feature preprocessing was performed using the training dataset only. To reduce redundancy between genomic and taxonomic features, SV features strongly correlated with species abundance features (Spearman |r| > 0.7) were removed. Remaining SV features showing high inter-feature correlation (Spearman |r| > 0.8) were further excluded.

Feature selection was performed using least absolute shrinkage and selection operator (LASSO) regression implemented in the glmnet package^40,41^. Ten-fold cross-validation was applied to determine optimal regularization parameters, and features were selected using both λ_1se_ and λ_min_ criteria. Separate feature selection procedures were performed for species-only models and combined species-plus-SV models.

Random forest classifiers were subsequently constructed using the ranger package with 1,000 trees and probability estimation enabled^42,43^. Model performance was evaluated using receiver operating characteristic (ROC) curves and area under the curve (AUC). Differences between paired ROC curves were assessed using DeLong’s test^44^ implemented in the pROC package^45^.

Model interpretation was performed using SHAP analysis^46^ implemented in the treeshap package. Feature importance was quantified using mean absolute SHAP values, and cross-cohort consistency of feature contributions was evaluated by comparing SHAP-based feature rankings between training and independent validation cohorts.

### Functional Annotation of SV-Associated Genes

To investigate the potential functional consequences of disease-associated structural variations, genes located within SV-associated genomic bins were identified based on the corresponding reference genome annotations. Only SV regions containing annotated protein-coding genes were selected for downstream functional characterization. Conserved protein domains and functional features of SV-associated genes were annotated using the NCBI Conserved Domain Database (CD-Search)^47^. Significant domain assignments, conserved catalytic residues, and predicted functional sites were obtained directly from the NCBI CD-Search output.

### Strain-Level Phylogenetic Analysis and Gene Distribution Analysis

To evaluate the strain-level distribution of SV-associated genes, complete chromosome-level genome assemblies were retrieved from the NCBI Assembly database for the corresponding bacterial species. Reference genome sequences were screened using BLASTN against SV-associated gene sequences to determine the presence or absence of the corresponding SV-associated genes across strains. Genome annotations were generated using Prokka^48^. Pan-genome analysis and core genome alignment generation were performed using Panaroo^49^ in strict cleaning mode with a 50% core gene threshold (--core_threshold 0.5). Phylogenetic reconstruction was performed using FastTree based on the core genome alignment using the Generalized Time-Reversible (GTR) nucleotide substitution model (-nt -gtr)^50^. The presence or absence patterns of SV-associated genes across strains were visualized together with the phylogenetic tree.

### Conserved Motif Identification and Binding Site Prediction

To investigate whether the SV-associated XRE-family regulator mediates autoregulatory feedback through conserved promoter binding, homologous protein sequences were identified and the upstream regulatory regions of their corresponding genes were analyzed for conserved DNA-binding motifs. Because XRE-family transcriptional regulators frequently regulate their own expression through promoter binding, homologous protein sequences of the target regulator were identified using BLASTP against related bacterial genomes. For each homologous protein, the corresponding upstream promoter regions (500 bp) were extracted from genome sequences based on the annotated gene coordinates.

Conserved DNA-binding motifs were identified using the MEME Suite^51^. MEME analysis was performed in DNA mode with the following parameters: zero or one occurrence per sequence (zoops) model, five candidate motifs, motif widths ranging from 12 to 20 bp, classic objective function, reverse-complement strand search, and zero-order Markov background model. The identified conserved motifs were subsequently scanned against the complete reference genome sequence using FIMO with a significance threshold of P < 1 × 10^-4^. Motif occurrences were mapped to genomic coordinates to identify potential regulatory binding regions. Motif significance was evaluated based on FIMO-adjusted P values, and candidate regulatory loci were further characterized through genomic annotation and functional interpretation.

### Protein–DNA Complex Structural Prediction

To investigate the potential molecular interaction between SV-associated regulatory proteins and their target DNA sequences, protein–DNA complex structures were predicted using AlphaFold 3^52^. Structural models were generated for wild-type and motif-mutated DNA sequences corresponding to the predicted binding regions. Predicted complex confidence was evaluated using AlphaFold3-derived structural confidence metrics, including predicted template modeling score (pTM) and interface predicted template modeling score (ipTM). Structural comparisons were performed to assess potential effects of SV-associated sequence variation on protein–DNA interactions.

## Data Availability

Whole-genome shotgun sequencing data have been deposited in the Genome Sequence Archive (GSA) at the National Genomics Data Center (accession numbers CRA004348 and PRJCA000337) and the European Bioinformatics Institute (EBI) database (accession number PRJEB6997).

## Abbreviations

RA: rheumatoid arthritis
ACPA: anti-citrullinated protein antibody
FMT: fecal microbiota transplantation
SV: structural variant
dSV: deletion structural variant
vSV: variable structural variant
OA: osteoarthritis
ESR: erythrocyte sedimentation rate
CRP: C-reactive protein
DMARD: disease-modifying antirheumatic drug
GSA: Genome Sequence Archive
EBI: European Bioinformatics Institute
ICRA: Iterative Coverage-based Read Assignment
TSS: total sum scaling
NMDS: non-metric multidimensional scaling
PCoA: principal coordinates analysis
PERMANOVA: permutational multivariate analysis of variance
QL: quasi-likelihood
TMM: trimmed mean of M-values
REML: restricted maximum likelihood
MH: Mantel–Haenszel
SMD: standardized mean difference
OR: odds ratio
LASSO: least absolute shrinkage and selection operator
ROC: receiver operating characteristic
AUC: area under the curve
SHAP: SHapley Additive exPlanations
CDD: Conserved Domain Database
XRE: Xenobiotic Response Element
TSS: transcription start site
SCFA: short-chain fatty acid
pTM: predicted template modeling score
ipTM: interface predicted template modeling score

## Ethics approval and consent to participate

Not applicable.

## Consent for publication

Not applicable.

## Statement

During the preparation of this work the authors used Gemini 3.2 in order to refine the English language. After using this tool, the authors reviewed and edited the content as needed and take full responsibility for the content of the published article.

## Authors’ contributions

Y.O., S.Z., J.W., Y.L., and H.L. contributed equally as co-first authors. Y.O. conceptualized the study, designed the methodology, and wrote the original manuscript draft. S.Z. was responsible for validation, laboratory investigation, and executing the biophysical surface plasmon resonance (SPR) assays. J.W. performed the cross-cohort meta-analysis, led data curation, and generated the primary data visualizations. Y.L. conducted the machine learning modeling, executed the genetics-related bioinformatics pipelines, and performed formal statistical analyses. H.L. contributed to methodology development, bioinformatics workflows, and computational AlphaFold modeling.

Y.H., S.L., Y.X., H.P., and Z.W. contributed to the clinical interpretation of the genomic findings, provided translational medicine insights, and participated in the conceptual design of the study. Q.X. contributed to big data platform management, multi-cohort dataset infrastructure, and preliminary data processing. L.S. and H.Z. served as co-corresponding authors, jointly supervising the research project. They acquired funding, provided overall scientific supervision, and were responsible for manuscript review, editing, and final approval. All authors reviewed the manuscript drafts and approved the final version for submission.

## Acknowledgements

We would like to express our gratitude to the researchers and consortia that made their data publicly available, enabling this work. We are also deeply grateful to the Medical Big Data Analysis Platform at Nanjing Drum Tower Hospital for providing the computational infrastructure and technical support. Additionally, we would like to thank all staff members of the Department of Rheumatology and Immunology at Nanjing Drum Tower Hospital for their continuous support and valuable insights throughout this study.

---

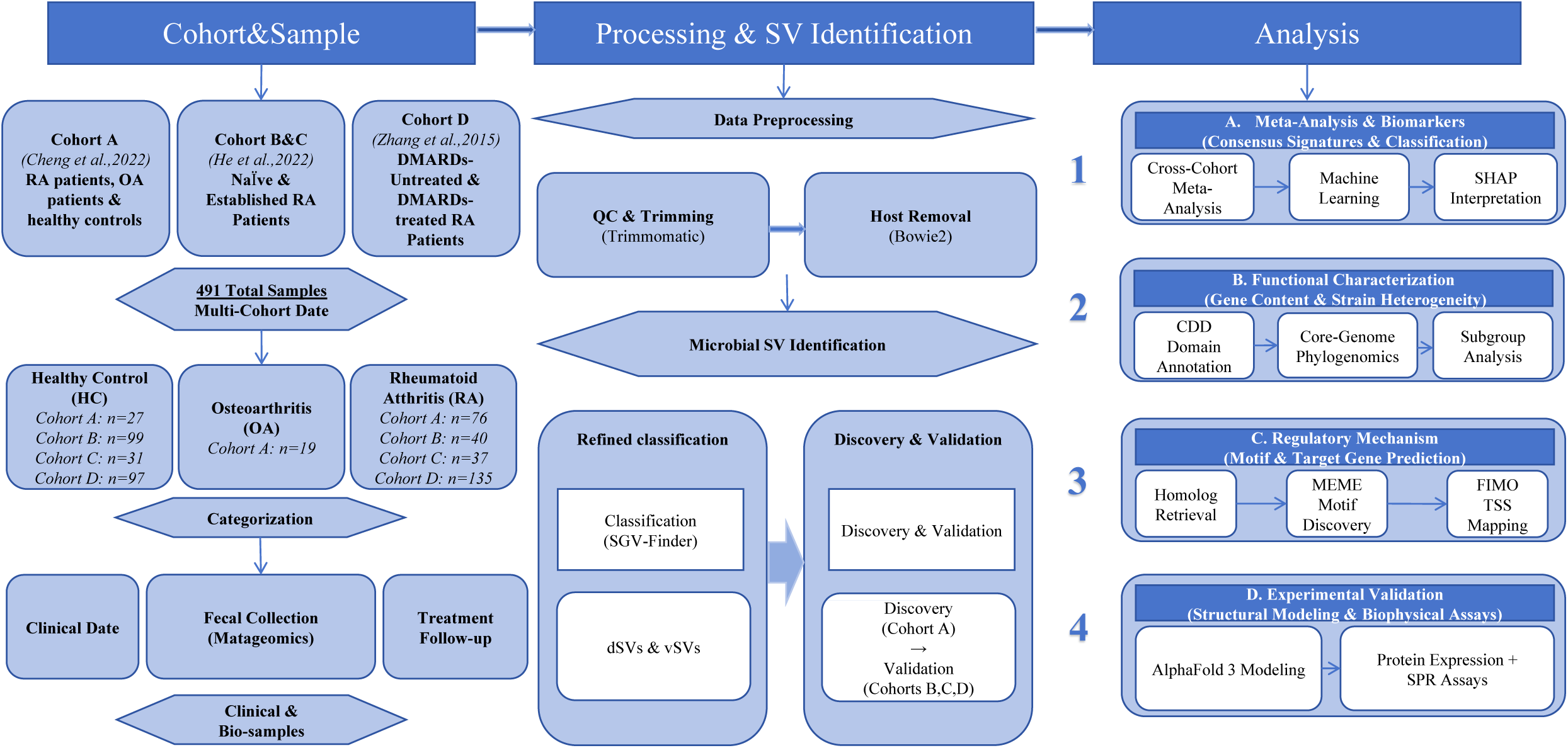

## Supplementary Figure Legends

**Figure S1. Gut microbiota diversity and community composition analyses across health and disease cohorts in Cohort A.**

(A–D) Alpha diversity comparison among Healthy Controls (HC), Osteoarthritis (OA), and Rheumatoid Arthritis (RA) groups evaluated by Shannon Index (A, p = 0.089), Simpson Index (B, p = 0.096), Observed Richness (C, p = 0.001), and Pielou’s Evenness (D, p = 0.178). (E, F) Beta diversity ordination analyzed by Non-metric Multidimensional Scaling (NMDS; Stress = 0.198) (E) and Principal Coordinate Analysis (PCoA; PCoA1 = 24.9%, PCoA2 = 17.9%) (F) based on Bray-Curtis dissimilarity, with confidence ellipses representing distinct clinical groups.

**Figure S2. Alpha and beta diversity evaluation of gut microbial communities in Cohort B.**

(A–D) Alpha diversity metrics comparing Healthy Controls (HC) and Rheumatoid Arthritis (RA) patients across Shannon Index (A, p = 0.871), Simpson Index (B, p = 0.947), Observed Richness (C, p = 0.539), and Pielou’s Evenness (D, p = 0.880). (E, F) Community-level beta diversity ordinations illustrated by NMDS (E) and PCoA (F; PCoA1 = 22.9%, PCoA2 = 18.4%) plots based on Bray-Curtis distance metrics.

**Figure S3. Gut microbial alpha and beta diversity assessment in Cohort C.**

(A–D) Comparison of alpha diversity indices between Healthy Controls (HC) and Rheumatoid Arthritis (RA) individuals for Shannon Index (A, p = 0.463), Simpson Index (B, p = 0.345), Observed Richness (C, p = 0.649), and Pielou’s Evenness (D, p = 0.405). (E, F) Beta diversity distributions visualized via NMDS (E) and PCoA (F; PCoA1 = 21.5%, PCoA2 = 16.6%) models.

**Figure S4. Gut microbiota diversity profiling according to treatment status in Cohort D.**

(A–D) Alpha diversity comparison across Control, Treated RA, and Untreated RA groups measured by Shannon Index (A, p = 0.430), Simpson Index (B, p = 0.448), Observed Richness (C, p = 0.236), and Pielou’s Evenness (D, p = 0.575). (E, F) Beta diversity ordinations demonstrated through NMDS (E; Stress = 0.255) and PCoA (F; PCoA1 = 18.8%, PCoA2 = 10.4%) profiles.

**Figure S5. Cross-cohort validation of quantile-scaled SHAP summaries for the consistently prioritized union set of structural variants (SVs).**

SHAP value distribution plots ranking key microbial SV features in the Training Cohort (Cohorts B+C) (A) and validated in the Independent Test Cohort (Cohort D) (B) (union of top 20 ranked features in Training Cohort B+C or Independent Test Cohort D). Feature values are color-coded from low (blue) to high (red) according to percentile rank, demonstrating consistent directionality and predictive importance across cohorts.

**Figure S6. Forest plots of meta-analyzed odds ratios for consistently prioritized deletion structural variants (dSVs) across cohorts.**

Forest plots displaying meta-analyzed odds ratios and 95% CIs for consistently prioritized deletion SV loci across Cohorts A–D, including *Oscillibacter* sp.: 557_558;1131_1132 (A), *Bacteroides xylanisolvens*: 5601_5602 (B), *Agathobacter rectalis*: 662_663 (C), *Faecalibacterium prausnitzii*: 537_538 (D), and *Faecalibacterium prausnitzii*: 781_783 (E). Summary Odds Ratios (ORs), 95% Confidence Intervals (CIs), and heterogeneity statistics (I² and Cochrane’s Q test p-values) are indicated.

**Figure S7. Meta-analysis of consistently prioritized variable structural variants (vSVs) across multi-center cohorts.**

Forest plots displaying meta-analyzed odds ratios and 95% CIs for consistently prioritized variable SV loci across Cohorts A–D, including Bacteroides fragilis: 4781_4788 (A), *Parabacteroides merdae*: 782_783;2923_2924 (B), *Bacteroides uniformis*: 3880_3883 (C), *Agathobacter rectalis*: 3036_3037 (D), *Agathobacter rectalis*: 3102_3105;3105_3106 (E), *Ruminococcus bromii*: 97_103 (F), and *Alistipes shahii*: 2252_2255;2255_2257;2257_2259;2259_2262;2262_2264;2265_2266 (G).

**Figure S8. Leave-one-out (LOO) sensitivity analyses for consistently prioritized deletion structural variants (dSVs).**

Sensitivity plots assessing the stability of pooled effect sizes after sequentially omitting each cohort for *Oscillibacter* sp.: 557_558;1131_1132 (A), *Bacteroides xylanisolvens*: 5601_5602 (B), *Agathobacter rectalis*: 662_663 (C), *Faecalibacterium prausnitzii*: 537_538 (D), and *Faecalibacterium prausnitzii*: 781_783 (E).

**Figure S9. Leave-one-out (LOO) sensitivity analyses for consistently prioritized variable structural variants (vSVs).**

Sensitivity plots assessing the stability of pooled effect sizes after sequentially omitting each cohort for Bacteroides fragilis: 4781_4788 (A), *Parabacteroides merdae:* 782_783;2923_2924 (B), *Bacteroides uniformis*: 3880_3883 (C), *Agathobacter rectalis*: 3036_3037 (D), *Agathobacter rectalis*: 3102_3105;3105_3106 (E), and *Ruminococcus bromii*: 97_103 (F), and *Alistipes shahii*: 2252_2255;2255_2257;2257_2259;2259_2262;2262_2264;2265_2266 (G).

**Figure S10. Comparative prevalence analysis of significant deletion SVs across disease and treatment subgroups.**

Proportions of structural variant presence (blue) and deletion (orange) for *Faecalibacterium prausnitzii*: 781_783 compared between Osteoarthritis (OA) vs. Rheumatoid Arthritis (RA) controls (p = 1) (A) and between Treated vs. Untreated RA patients (p = 1) (B).

**Figure S11. Functional domain annotation and catalytic site mapping of the retron-associated reverse transcriptase locus (*HMPREF9436_01829*) in *Faecalibacterium prausnitzii*: 781_783.**

(A) Domain significance analysis indicating the domain architecture of the deletion region, dominated by cd03487: RT_Bac_retron_II (-log10 E-value = 54.32) and NF038233: retron_St85_RT (-log10 E-value = 36.51). (B) Detailed residue-level site profiling illustrating conserved catalytic centers, including putative active sites, NTP-binding sites, and nucleic acid-binding sites within the retron reverse transcriptase protein.

**Figure S12. Phylogenomic distribution and structural presence/absence of the retron reverse transcriptase locus (*HMPREF9436_01829*) across *Faecalibacterium prausnitzii* strains.**

Strain-level phylogenetic tree constructed from *Faecalibacterium prausnitzii* isolates, displaying the lineage-specific presence (filled blue) and structural deletion (unfilled red) of the target retron-associated reverse transcriptase locus across distinct bacterial strains.

**Figure S13. De novo motif discovery of the conserved promoter binding sequence for EUR_RS03225.**

Sequence logo of the top consensus DNA-binding motif (top motif E-value = 7.20E-88) identified by MEME analysis across the 500-bp upstream promoter regions of EUR_RS03225 homologous genes obtained via BLASTP. The sequence demonstrates a characteristic palindromic (inverted repeat) structure corresponding to the dimeric DNA-binding mode of XRE-family transcriptional regulators.

**Figure S14. Domain annotation and genomic architecture of the predicted downstream short-chain fatty acid metabolic locus (*EUR_RS01730*).**

Conserved domain analysis of the metabolic gene cluster (*EUR_RS01730*) adjacent to the EUR_RS03225 binding site, highlighting key acetyl-CoA acetyltransferase domain elements including TIGR01930: AcCoA-C-Actrans, PRK05790, PaaJ, and thiolase (-log10 E-value = 300.0) involved in acetyl-CoA condensation, a key step in microbial butyrate biosynthesis.

**Figure S15. 3D structural modeling and molecular docking of the EUR_RS03225 regulator dimer with predicted target DNA sequences and their mutant counterparts.**

Predicted 3D structural complexes of the SV-associated transcriptional regulator dimer (EUR_RS03225) bound to target DNA sequences. Predicted binding interfaces are shown for wild-type sequences (ipTM = 0.74, pTM = 0.79 for *EUR_RS03225*_WT; ipTM = 0.72, pTM = 0.77 for *EUR_RS01730*_WT), alongside structural perturbations and reduced binding affinities induced by sequence mutations (ipTM = 0.67 and 0.69, respectively).

