## supplementary figures for "Gut microbial structural variation improves disease discrimination and reveals a strain-level regulatory mechanism in rheumatoid arthritis"

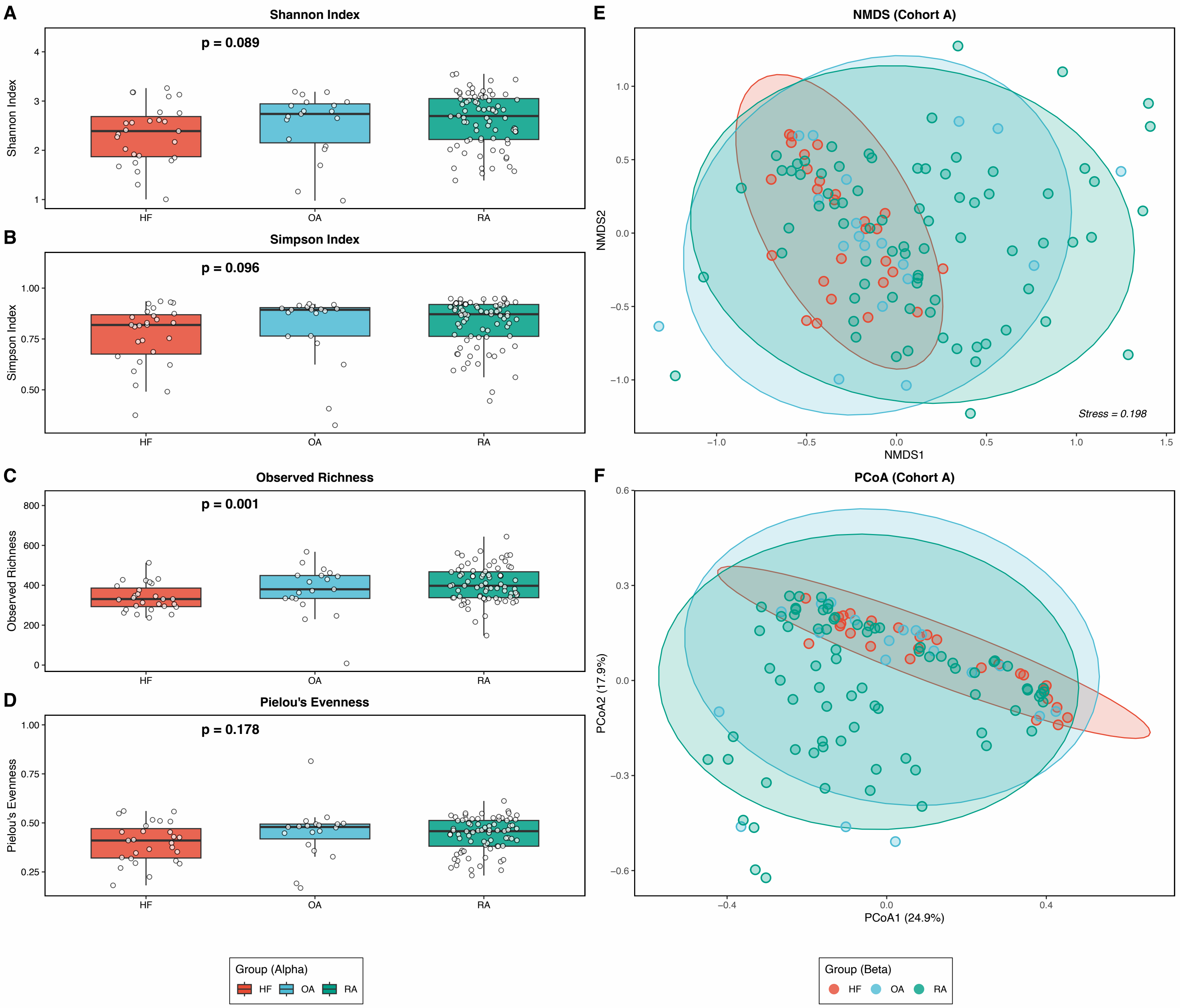
A
Shannon Index
p = 0.089
Shannon Index
HF
OA
RA
B
Simpson Index
p = 0.096
Simpson Index
C
Observed Richness
p = 0.001
Observed Richness
D
Pielou's Evenness
p = 0.178
Pielou's Evenness
E
NMDS (Cohort A)
NMDS2
NMDS1
Stress = 0.198
F
PCoA (Cohort A)
PCoA2 (17.9%)
PCoA1 (24.9%)
Group (Alpha)
HF OA RA
Group (Beta)
HF OA RA

Figure S1

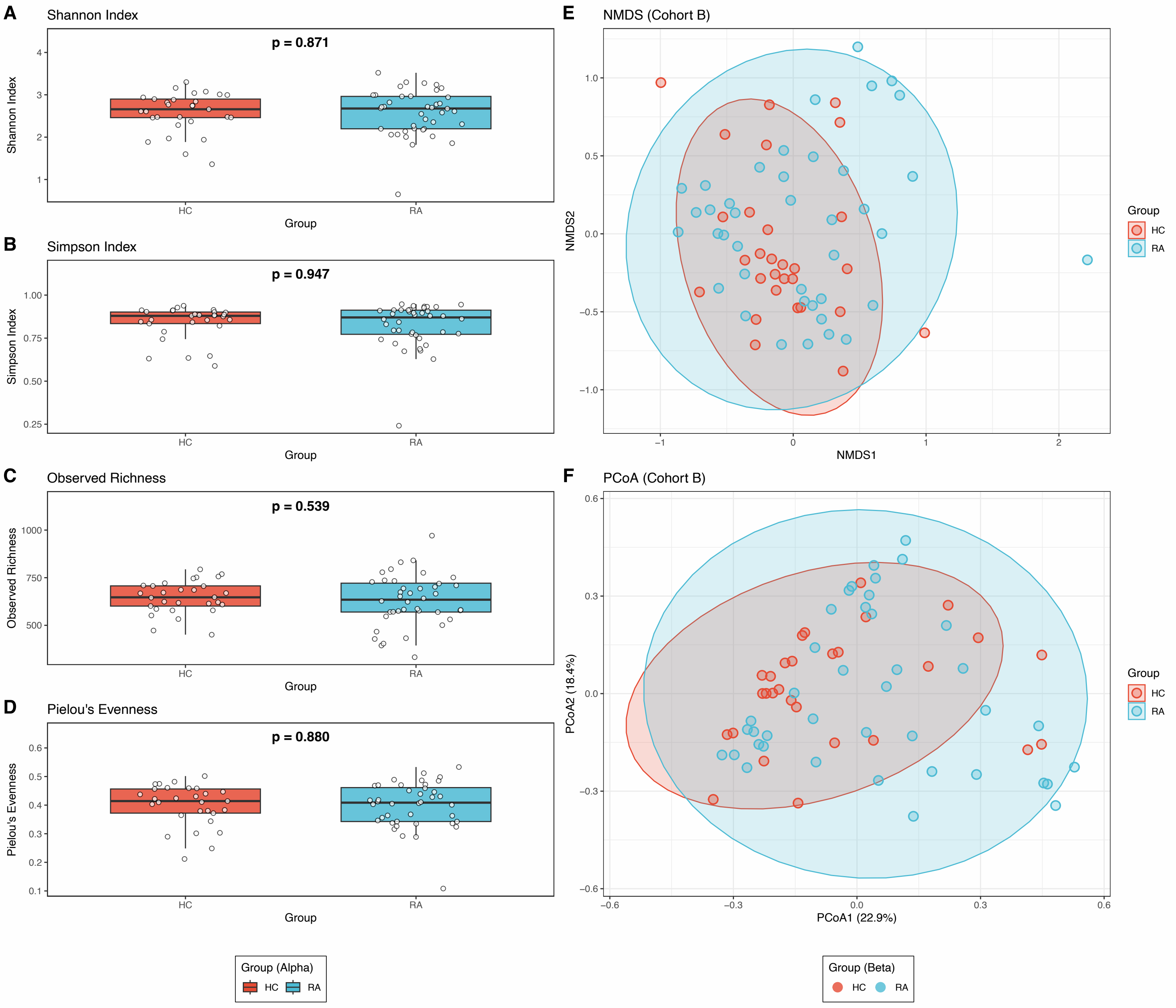
A
Shannon Index
p = 0.871
Shannon Index
HC
RA
Group
B
Simpson Index
p = 0.947
Simpson Index
HC
RA
Group
C
Observed Richness
p = 0.539
Observed Richness
HC
RA
Group
D
Pielou's Evenness
p = 0.880
Pielou's Evenness
HC
RA
Group
E
NMDS (Cohort B)
NMDS2
NMDS1
Group
HC
RA
F
PCoA (Cohort B)
PCoA2 (18.4%)
PCoA1 (22.9%)
Group
HC
RA
Group (Alpha)
HC
RA
Group (Beta)
HC
RA

Figure S2

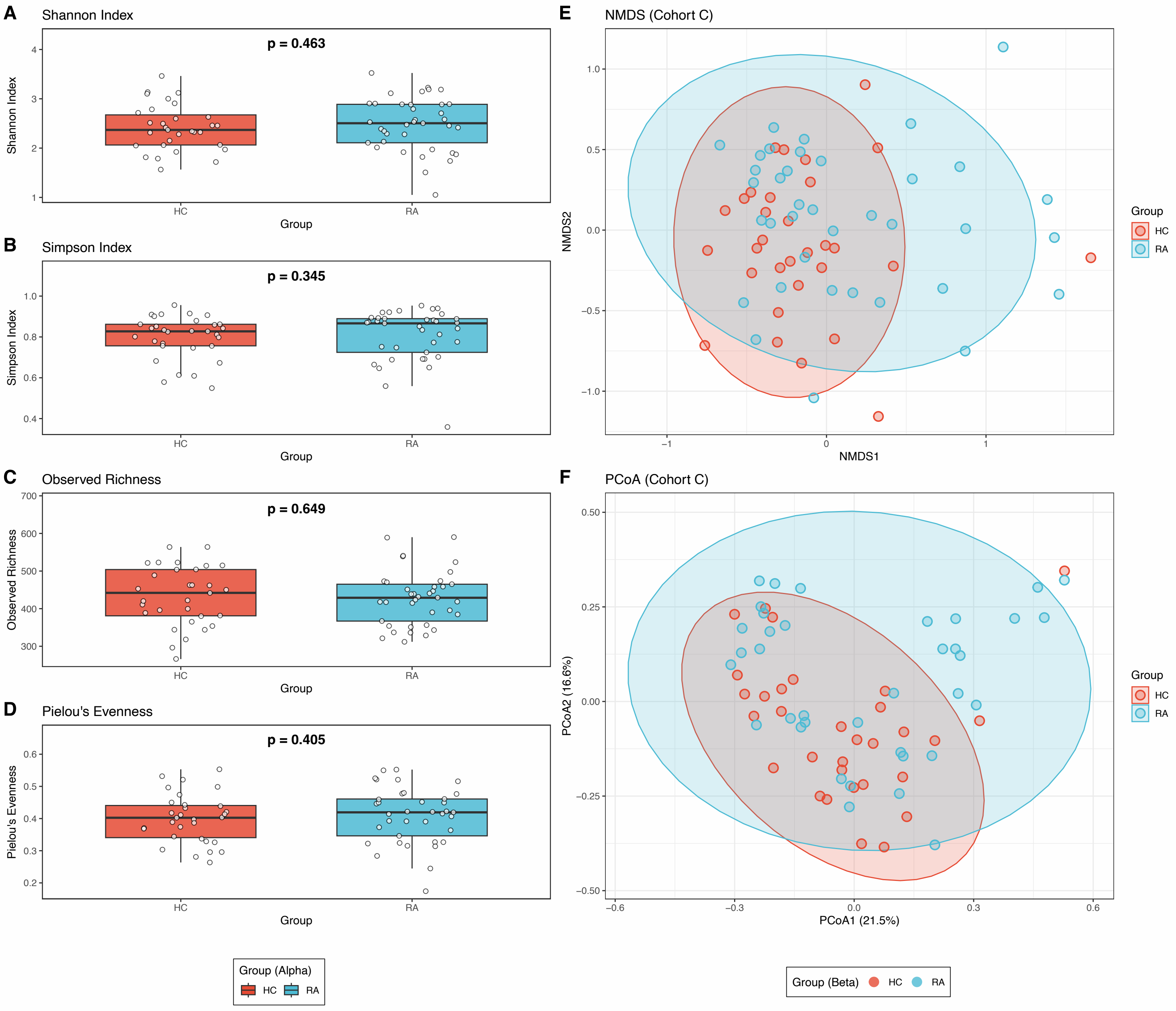

A
Shannon Index
p = 0.463
Shannon Index
Group
HC
RA
B
Simpson Index
p = 0.345
Simpson Index
C
Observed Richness
p = 0.649
Observed Richness
D
Pielou's Evenness
p = 0.405
Pielou's Evenness
Group (Alpha)
E
NMDS (Cohort C)
NMDS1
NMDS2
F
PCoA (Cohort C)
PCoA1 (21.5%)
PCoA2 (16.6%)
Group (Beta)


Figure S3

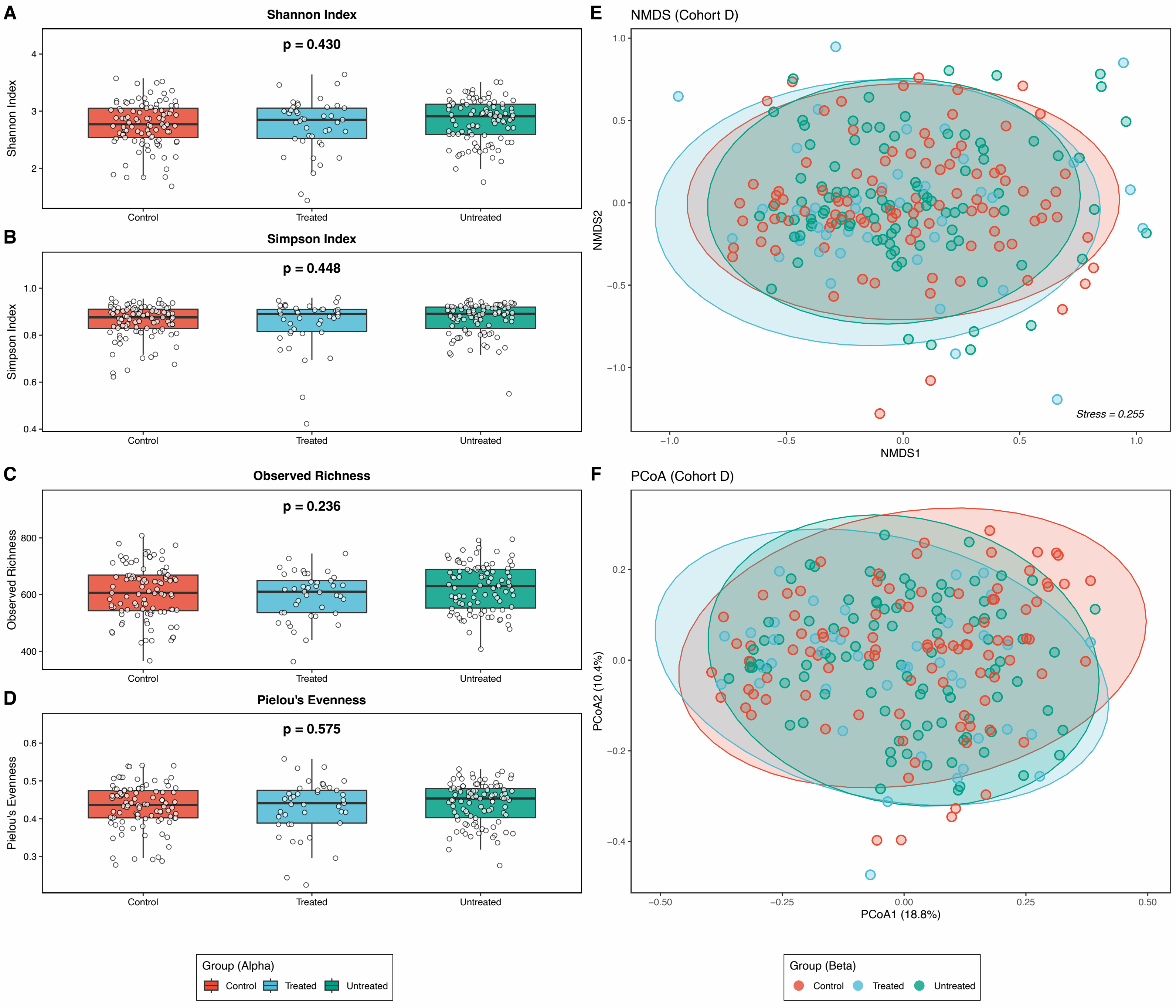

A
Shannon Index
p = 0.430
Shannon Index
Control
Treated
Untreated
B
Simpson Index
p = 0.448
Simpson Index
C
Observed Richness
p = 0.236
Observed Richness
D
Pielou's Evenness
p = 0.575
Pielou's Evenness
E
NMDS (Cohort D)
NMDS2
NMDS1
Stress = 0.255
F
PCoA (Cohort D)
PCoA2 (10.4%)
PCoA1 (18.8%)
Group (Alpha)
Control
Treated
Untreated
Group (Beta)
Control
Treated
Untreated


Figure S4

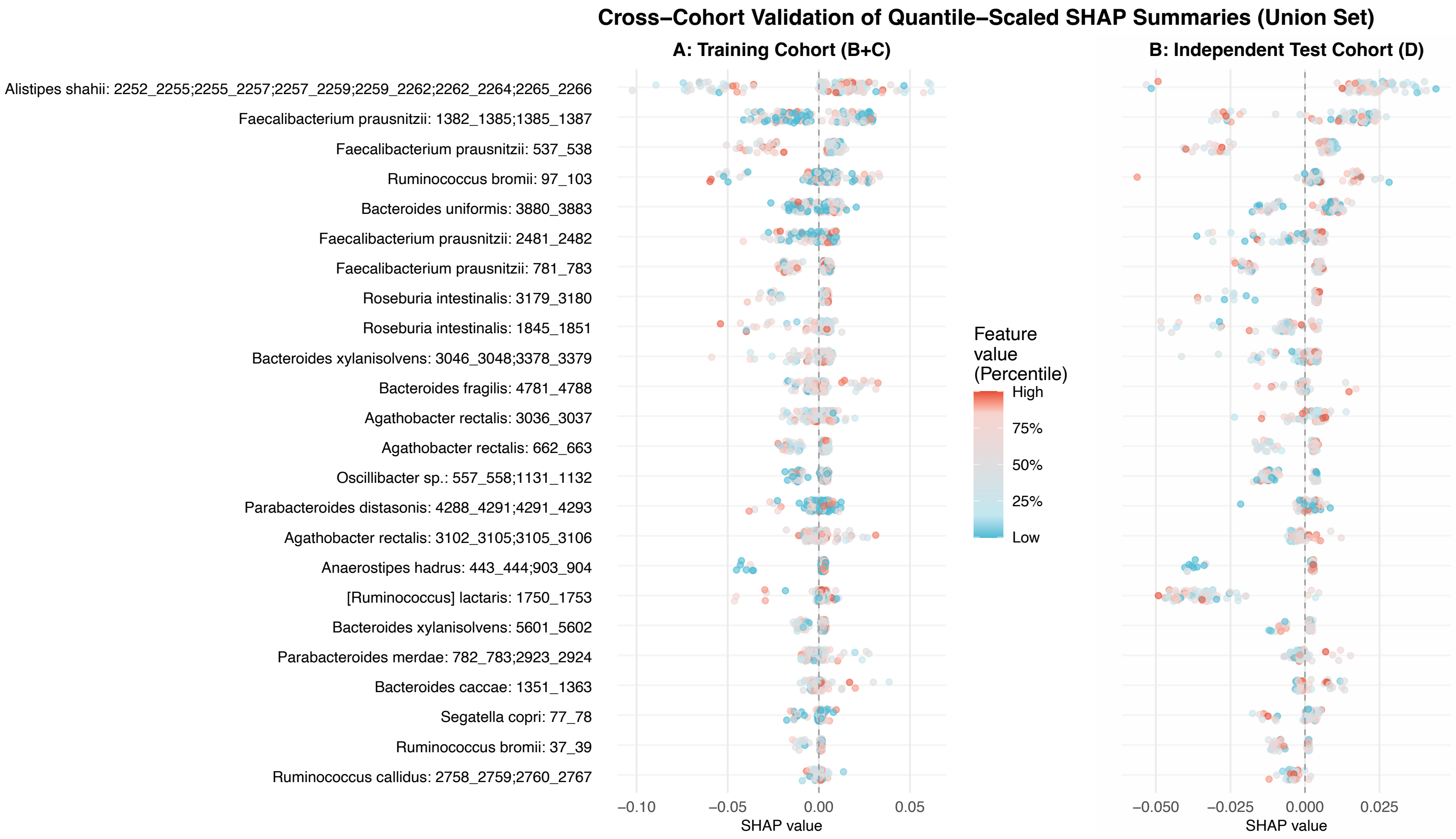

Cross–Cohort Validation of Quantile–Scaled SHAP Summaries (Union Set)
A: Training Cohort (B+C)
B: Independent Test Cohort (D)
Alistipes shahii: 2252_2255;2255_2257;2257_2259;2259_2262;2262_2264;2265_2266
Faecalibacterium prausnitzii: 1382_1385;1385_1387
Faecalibacterium prausnitzii: 537_538
Ruminococcus bromii: 97_103
Bacteroides uniformis: 3880_3883
Faecalibacterium prausnitzii: 2481_2482
Faecalibacterium prausnitzii: 781_783
Roseburia intestinalis: 3179_3180
Roseburia intestinalis: 1845_1851
Bacteroides xylanisolvens: 3046_3048;3378_3379
Bacteroides fragilis: 4781_4788
Agathobacter rectalis: 3036_3037
Agathobacter rectalis: 662_663
Oscillibacter sp.: 557_558;1131_1132
Parabacteroides distasonis: 4288_4291;4291_4293
Agathobacter rectalis: 3102_3105;3105_3106
Anaerostipes hadrus: 443_444;903_904
[Ruminococcus] lactaris: 1750_1753
Bacteroides xylanisolvens: 5601_5602
Parabacteroides merdae: 782_783;2923_2924
Bacteroides caccae: 1351_1363
Segatella copri: 77_78
Ruminococcus bromii: 37_39
Ruminococcus callidus: 2758_2759;2760_2767
−0.10
−0.05
0.00
0.05
SHAP value
−0.050
−0.025
0.000
0.025
SHAP value
Feature value (Percentile)
High
75%
50%
25%
Low


Figure S5

**A. Oscillibacter sp.: 557_558;1131_1132**

| Study | Comparison (Fractions) |
|---|---|
| Cohort D | 0.40 [0.11; 1.40] |
| Cohort A | 1.33 [0.10; 17.28] |
| Cohort B | 0.36 [0.09; 1.51] |
| Cohort C | 0.44 [0.09; 2.15] |
| Total | 0.44 [0.20; 0.96] |

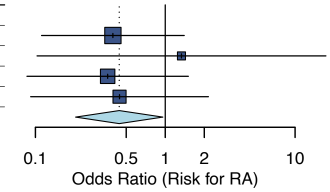


Heterogeneity: $\chi^2_3 = 0.82$ ($P$ = .84), $I^2 = 0.0\%$

**B. Bacteroides xylanisolvens: 5601_5602**

| Study | Comparison (Fractions) |
|---|---|
| Cohort D | 1.31 [0.34; 5.05] |
| Cohort A | 1.19 [0.20; 6.99] |
| Cohort B | 2.80 [0.62; 12.66] |
| Cohort C | 5.40 [1.12; 26.04] |
| Total | 2.18 [1.02; 4.69] |

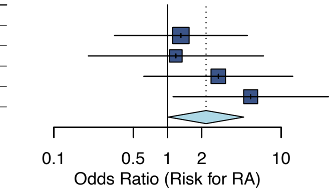


Heterogeneity: $\chi^2_3 = 2.38$ ($P$ = .50), $I^2 = 0.0\%$

**C. Agathobacter rectalis: 662_663**

| Study | Comparison (Fractions) |
|---|---|
| Cohort D | 0.56 [0.15; 2.02] |
| Cohort A | 0.41 [0.12; 1.40] |
| Cohort B | 0.41 [0.11; 1.58] |
| Cohort C | 0.20 [0.04; 1.12] |
| Total | 0.40 [0.20; 0.79] |

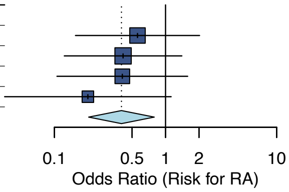


Heterogeneity: $\chi^2_3 = 0.89$ ($P$ = .83), $I^2 = 0.0\%$

**D. Faecalibacterium prausnitzii:537_538**

| Study | Comparison (Fractions) |
|---|---|
| Cohort D | 0.96 [0.32; 2.91] |
| Cohort A | 0.54 [0.18; 1.57] |
| Cohort B | 0.43 [0.12; 1.47] |
| Cohort C | 0.09 [0.01; 0.82] |
| Total | 0.53 [0.28; 0.99] |

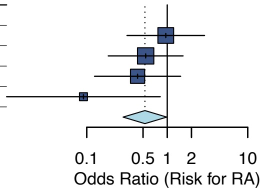


Heterogeneity: $\chi^2_3 = 3.68$ ($P$ = .30), $I^2 = 18.6\%$

**E. Faecalibacterium prausnitzii: 781_783**

| Study | Comparison (Fractions) |
|---|---|
| Cohort D | 0.41 [0.12; 1.39] |
| Cohort A | 0.49 [0.17; 1.44] |
| Cohort B | 0.35 [0.10; 1.20] |
| Cohort C | 0.47 [0.10; 2.17] |
| Total | 0.43 [0.23; 0.79] |

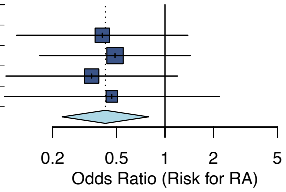


Heterogeneity: $\chi^2_3 = 0.18$ ($P$ = .98), $I^2 = 0.0\%$

Figure S6

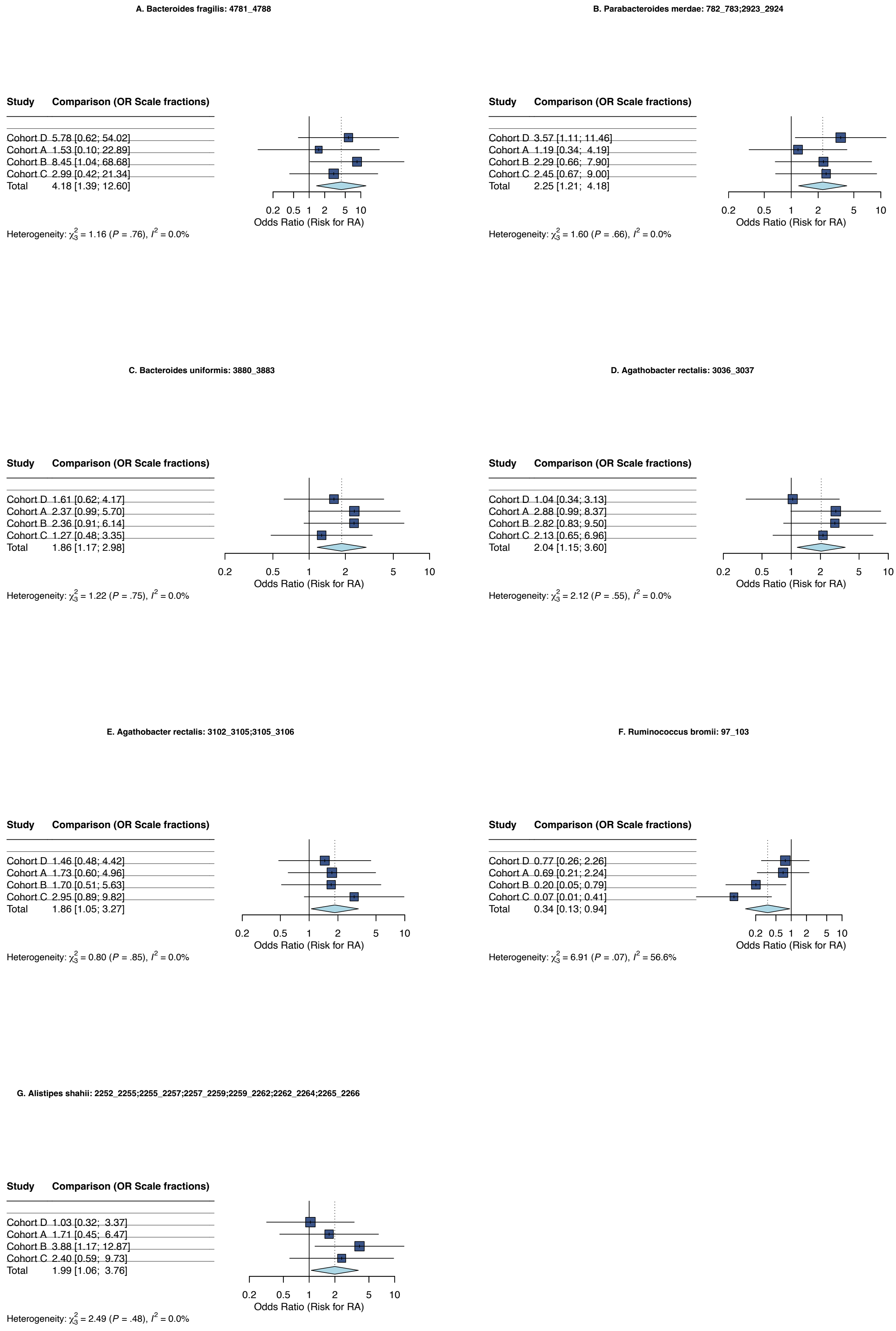

A. Bacteroides fragilis: 4781_4788
Study Comparison (OR Scale fractions)
Cohort D 5.78 [0.62; 54.02]
Cohort A 1.53 [0.10; 22.89]
Cohort B 8.45 [1.04; 68.68]
Cohort C 2.99 [0.42; 21.34]
Total 4.18 [1.39; 12.60]
0.2 0.5 1 2 5 10
Odds Ratio (Risk for RA)
Heterogeneity: $\chi^2_3$ = 1.16 (P = .76), $I^2$ = 0.0%
B. Parabacteroides merdae: 782_783;2923_2924
Study Comparison (OR Scale fractions)
Cohort D 3.57 [1.11; 11.46]
Cohort A 1.19 [0.34; 4.19]
Cohort B 2.29 [0.66; 7.90]
Cohort C 2.45 [0.67; 9.00]
Total 2.25 [1.21; 4.18]
0.2 0.5 1 2 5 10
Odds Ratio (Risk for RA)
Heterogeneity: $\chi^2_3$ = 1.60 (P = .66), $I^2$ = 0.0%
C. Bacteroides uniformis: 3880_3883
Study Comparison (OR Scale fractions)
Cohort D 1.61 [0.62; 4.17]
Cohort A 2.37 [0.99; 5.70]
Cohort B 2.36 [0.91; 6.14]
Cohort C 1.27 [0.48; 3.35]
Total 1.86 [1.17; 2.98]
0.2 0.5 1 2 5 10
Odds Ratio (Risk for RA)
Heterogeneity: $\chi^2_3$ = 1.22 (P = .75), $I^2$ = 0.0%
D. Agathobacter rectalis: 3036_3037
Study Comparison (OR Scale fractions)
Cohort D 1.04 [0.34; 3.13]
Cohort A 2.88 [0.99; 8.37]
Cohort B 2.82 [0.83; 9.50]
Cohort C 2.13 [0.65; 6.96]
Total 2.04 [1.15; 3.60]
0.2 0.5 1 2 5 10
Odds Ratio (Risk for RA)
Heterogeneity: $\chi^2_3$ = 2.12 (P = .55), $I^2$ = 0.0%
E. Agathobacter rectalis: 3102_3105;3105_3106
Study Comparison (OR Scale fractions)
Cohort D 1.46 [0.48; 4.42]
Cohort A 1.73 [0.60; 4.96]
Cohort B 1.70 [0.51; 5.63]
Cohort C 2.95 [0.89; 9.82]
Total 1.86 [1.05; 3.27]
0.2 0.5 1 2 5 10
Odds Ratio (Risk for RA)
Heterogeneity: $\chi^2_3$ = 0.80 (P = .85), $I^2$ = 0.0%
F. Ruminococcus bromii: 97_103
Study Comparison (OR Scale fractions)
Cohort D 0.77 [0.26; 2.26]
Cohort A 0.69 [0.21; 2.24]
Cohort B 0.20 [0.05; 0.79]
Cohort C 0.07 [0.01; 0.41]
Total 0.34 [0.13; 0.94]
0.2 0.5 1 2 5 10
Odds Ratio (Risk for RA)
Heterogeneity: $\chi^2_3$ = 6.91 (P = .07), $I^2$ = 56.6%
G. Alistipes shahii: 2252_2255;2255_2257;2257_2259;2259_2262;2262_2264;2265_2266
Study Comparison (OR Scale fractions)
Cohort D 1.03 [0.32; 3.37]
Cohort A 1.71 [0.45; 6.47]
Cohort B 3.88 [1.17; 12.87]
Cohort C 2.40 [0.59; 9.73]
Total 1.99 [1.06; 3.76]
0.2 0.5 1 2 5 10
Odds Ratio (Risk for RA)
Heterogeneity: $\chi^2_3$ = 2.49 (P = .48), $I^2$ = 0.0%


Figure S7

A. LOO Sensitivity: Oscillibacter sp.: 557_558;1131_1132

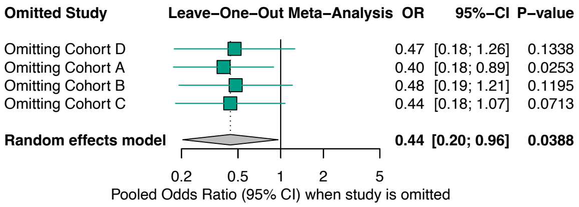


B. LOO Sensitivity: Bacteroides xylanisolvens: 5601_5602

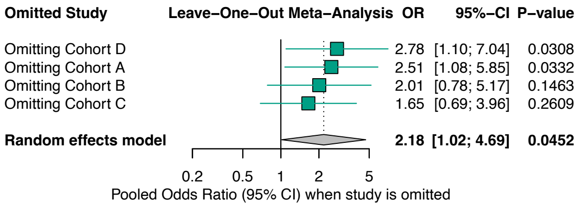


C. LOO Sensitivity: Agathobacter rectalis: 662_663

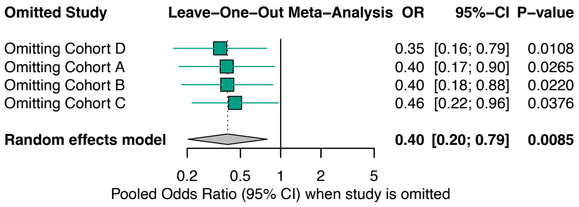


D. LOO Sensitivity: Faecalibacterium prausnitzii: 537_538

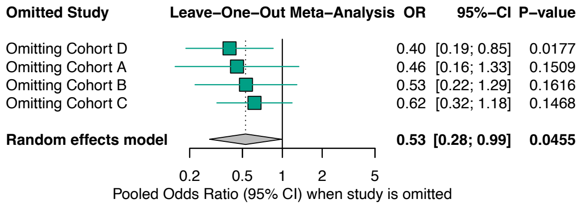


E. LOO Sensitivity:  Faecalibacterium prausnitzii: 781_783

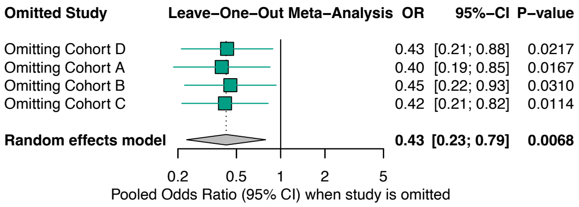


Figure S8

**A. LOO Sensitivity: Bacteroides fragilis: 4781_4788**

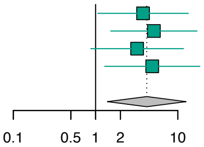


| Omitted Study | OR | 95% CI | P−value |
|---|---|---|---|
| Omitting Cohort D | 3.77 | [1.06; 13.39] | 0.0399 |
| Omitting Cohort A | 5.11 | [1.53; 17.09] | 0.0081 |
| Omitting Cohort B | 3.20 | [0.88; 11.69] | 0.0787 |
| Omitting Cohort C | 4.88 | [1.29; 18.48] | 0.0195 |
| **Random effects model** | **4.18** | **[1.39; 12.60]** | **0.0109** |

**B. LOO Sensitivity: Parabacteroides merdae: 782_783;2923_2924**

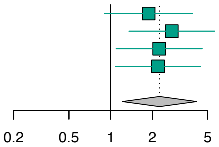


| Omitted Study | OR | 95% CI | P−value |
|---|---|---|---|
| Omitting Cohort D | 1.88 | [0.90; 3.90] | 0.0910 |
| Omitting Cohort A | 2.76 | [1.35; 5.61] | 0.0052 |
| Omitting Cohort B | 2.23 | [1.09; 4.57] | 0.0274 |
| Omitting Cohort C | 2.19 | [1.08; 4.43] | 0.0289 |
| **Random effects model** | **2.25** | **[1.21; 4.18]** | **0.0103** |

**C. LOO Sensitivity: Bacteroides uniformis: 3880_3883**

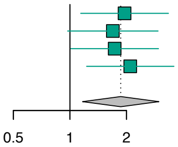


| Omitted Study | OR | 95% CI | P−value |
|---|---|---|---|
| Omitting Cohort D | 1.95 | [1.14; 3.34] | 0.0146 |
| Omitting Cohort A | 1.69 | [0.97; 2.94] | 0.0627 |
| Omitting Cohort B | 1.73 | [1.01; 2.96] | 0.0454 |
| Omitting Cohort C | 2.09 | [1.23; 3.58] | 0.0067 |
| **Random effects model** | **1.86** | **[1.17; 2.98]** | **0.0092** |

**D. LOO Sensitivity: Agathobacter rectalis: 3036_3037**

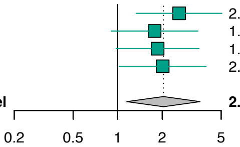


| Omitted Study | OR | 95% CI | P−value |
|---|---|---|---|
| Omitting Cohort D | 2.60 | [1.34; 5.05] | 0.0048 |
| Omitting Cohort A | 1.78 | [0.90; 3.48] | 0.0950 |
| Omitting Cohort B | 1.86 | [0.98; 3.55] | 0.0591 |
| Omitting Cohort C | 2.01 | [1.02; 3.96] | 0.0424 |
| **Random effects model** | **2.04** | **[1.15; 3.60]** | **0.0143** |

**E. LOO Sensitivity: Agathobacter rectalis: 3102_3105;3105_3106**

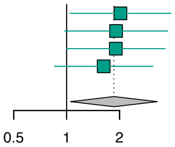


| Omitted Study | OR | 95% CI | P−value |
|---|---|---|---|
| Omitting Cohort D | 2.02 | [1.04; 3.92] | 0.0366 |
| Omitting Cohort A | 1.91 | [0.97; 3.75] | 0.0596 |
| Omitting Cohort B | 1.90 | [1.00; 3.63] | 0.0502 |
| Omitting Cohort C | 1.62 | [0.85; 3.09] | 0.1400 |
| **Random effects model** | **1.86** | **[1.05; 3.27]** | **0.0327** |

**F. LOO Sensitivity: Ruminococcus bromii: 97_103**

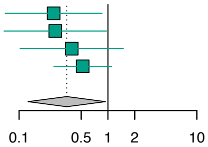


| Omitted Study | OR | 95% CI | P−value |
|---|---|---|---|
| Omitting Cohort D | 0.24 | [0.07; 0.86] | 0.0278 |
| Omitting Cohort A | 0.26 | [0.07; 0.96] | 0.0439 |
| Omitting Cohort B | 0.39 | [0.10; 1.49] | 0.1690 |
| Omitting Cohort C | 0.52 | [0.25; 1.10] | 0.0855 |
| **Random effects model** | **0.34** | **[0.13; 0.94]** | **0.0368** |

**G. LOO Sensitivity: Alistipes shahii: 2252_2255;2255_2257;2257_2259;2259_2262;2262_2264;2265_2266**

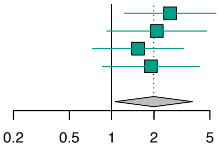


| Omitted Study | OR | 95% CI | P−value |
|---|---|---|---|
| Omitting Cohort D | 2.60 | [1.23; 5.51] | 0.0126 |
| Omitting Cohort A | 2.10 | [0.92; 4.76] | 0.0770 |
| Omitting Cohort B | 1.54 | [0.73; 3.25] | 0.2564 |
| Omitting Cohort C | 1.90 | [0.86; 4.22] | 0.1139 |
| **Random effects model** | **1.99** | **[1.06; 3.76]** | **0.0328** |

Figure S9

### Comparative Analysis of Significant dSVs

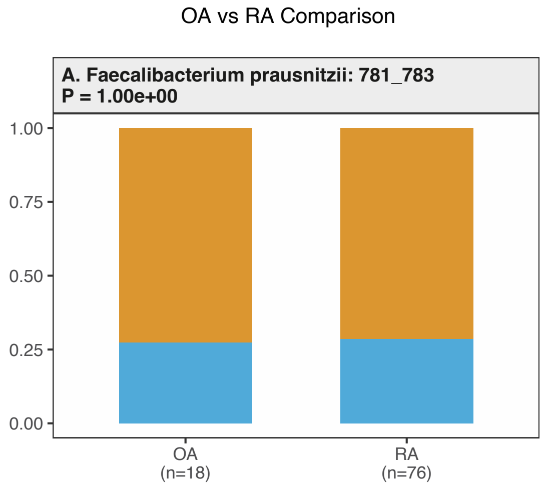


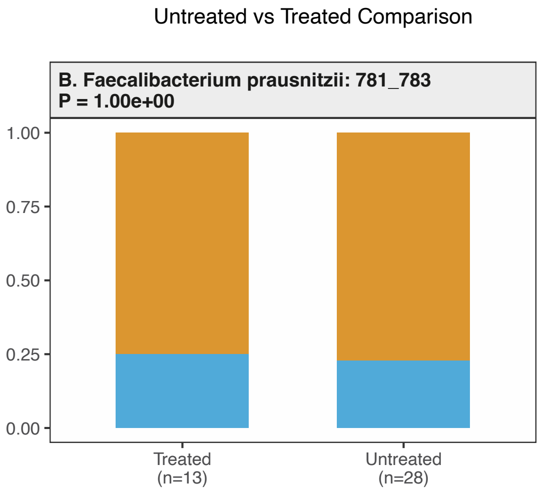


Figure S10

A

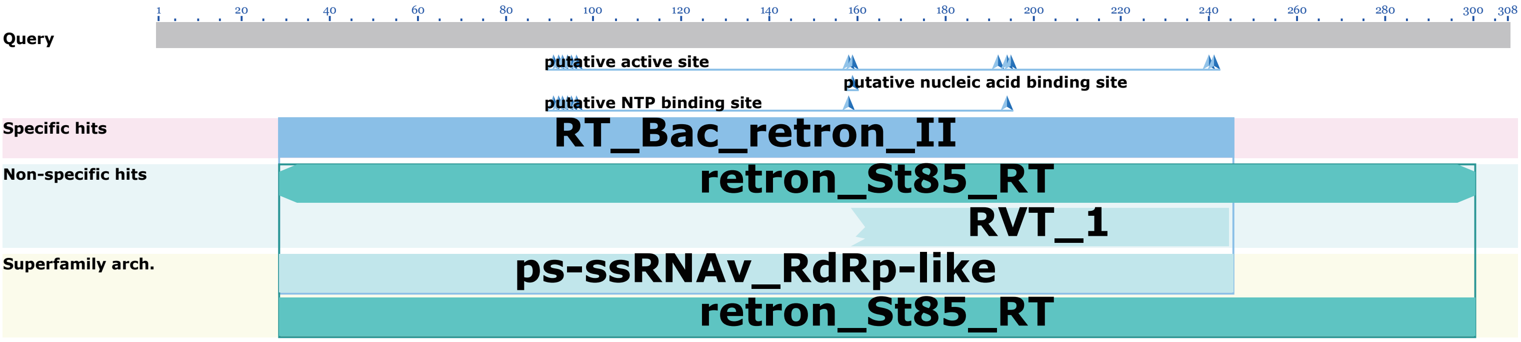


B

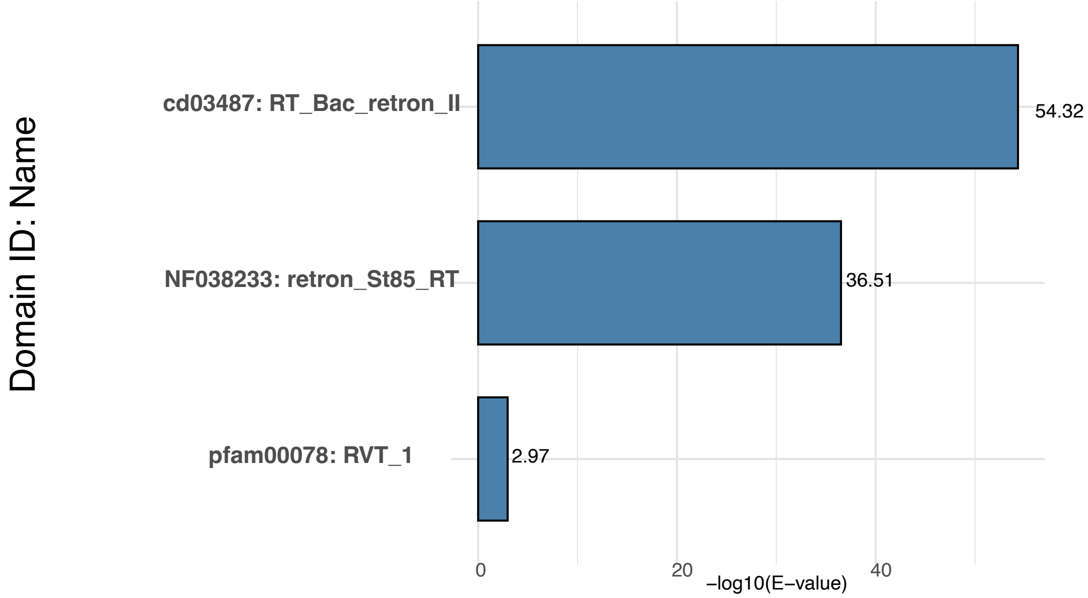


Figure S11

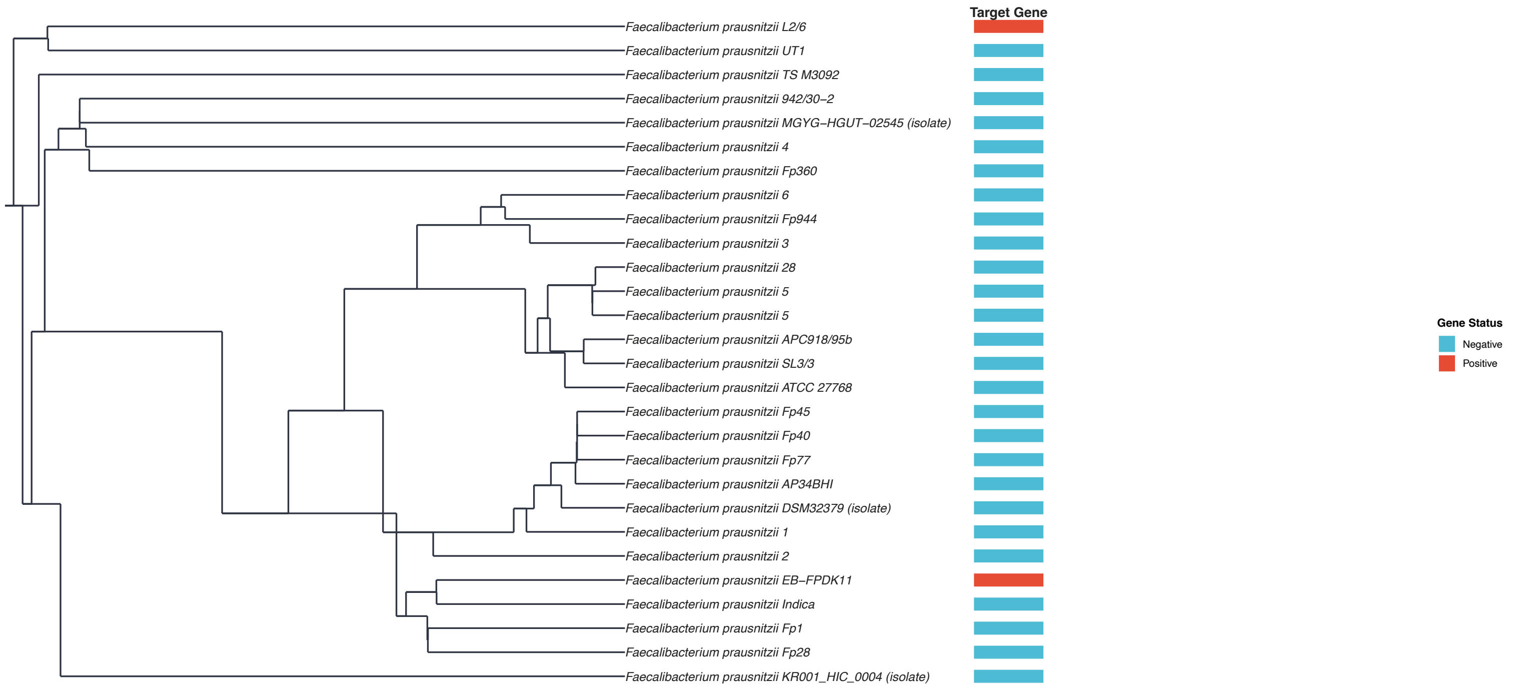

Target Gene
Faecalibacterium prausnitzii L2/6
Faecalibacterium prausnitzii UT1
Faecalibacterium prausnitzii TS M3092
Faecalibacterium prausnitzii 942/30−2
Faecalibacterium prausnitzii MGYG−HGUT−02545 (isolate)
Faecalibacterium prausnitzii 4
Faecalibacterium prausnitzii Fp360
Faecalibacterium prausnitzii 6
Faecalibacterium prausnitzii Fp944
Faecalibacterium prausnitzii 3
Faecalibacterium prausnitzii 28
Faecalibacterium prausnitzii 5
Faecalibacterium prausnitzii 5
Faecalibacterium prausnitzii APC918/95b
Faecalibacterium prausnitzii SL3/3
Faecalibacterium prausnitzii ATCC 27768
Faecalibacterium prausnitzii Fp45
Faecalibacterium prausnitzii Fp40
Faecalibacterium prausnitzii Fp77
Faecalibacterium prausnitzii AP34BHI
Faecalibacterium prausnitzii DSM32379 (isolate)
Faecalibacterium prausnitzii 1
Faecalibacterium prausnitzii 2
Faecalibacterium prausnitzii EB−FPDK11
Faecalibacterium prausnitzii Indica
Faecalibacterium prausnitzii Fp1
Faecalibacterium prausnitzii Fp28
Faecalibacterium prausnitzii KR001_HIC_0004 (isolate)
Gene Status
Negative
Positive


Figure S12

| No. | E-value | Sites | Width |
|---|---|---|---|
| 1 | 7.20E-88 | 22 | 20 |
| 2 | 4.60E-65 | 18 | 20 |
| 3 | 5.90E-49 | 22 | 20 |
| 4 | 5.40E-22 | 13 | 16 |
| 5 | 1.90E-08 | 18 | 12 |

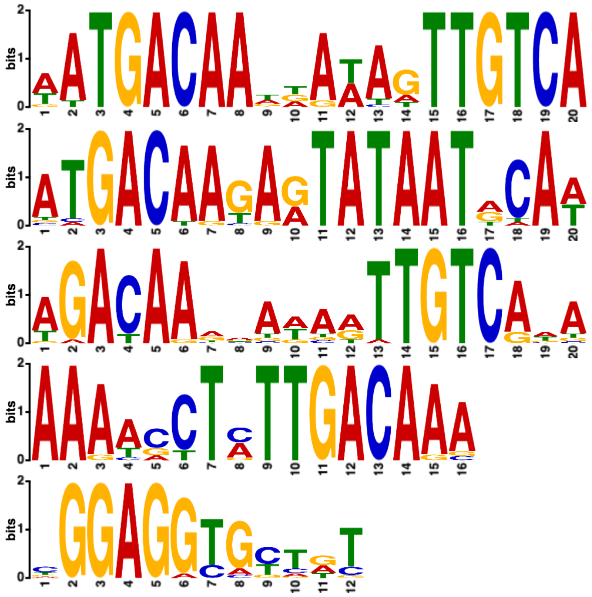

Figure S13

A

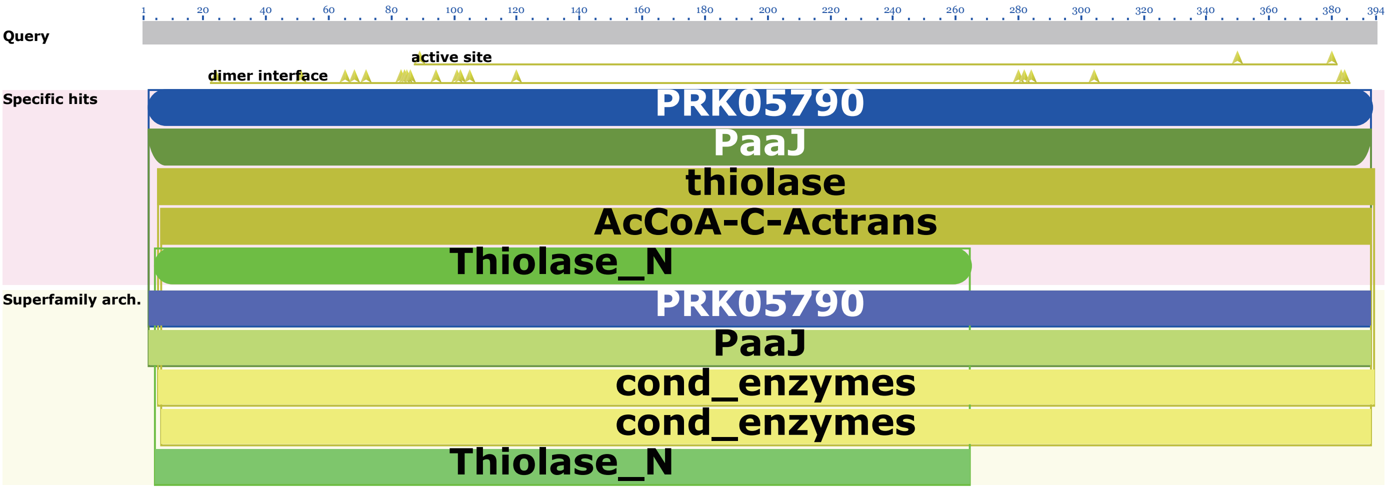


B

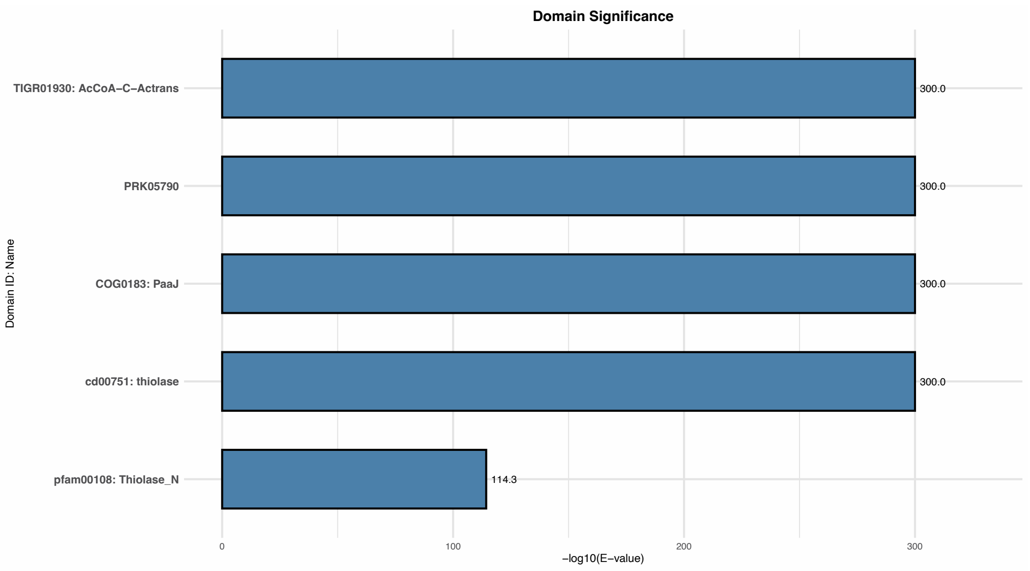


Figure S14

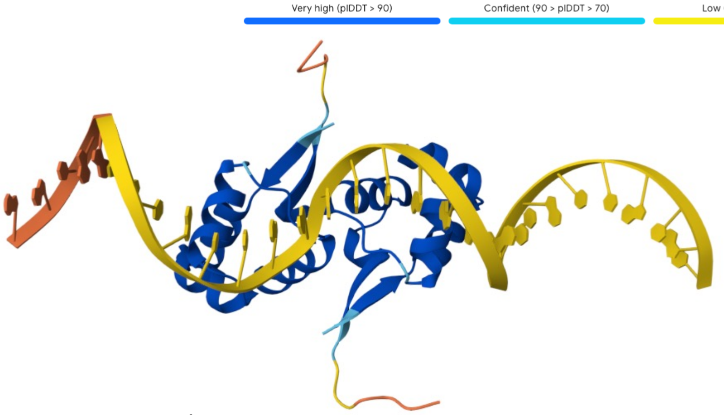


EUR_RS03225 dimer-
*EUR_RS03225*_WT:
ipTM = 0.74; pTM = 0.79

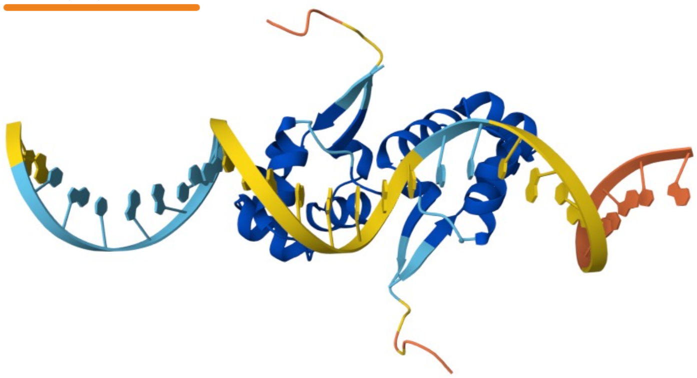

EUR_ RS03225 dimer-
*EUR_RS01730*_WT:
ipTM = 0.72; pTM = 0.77

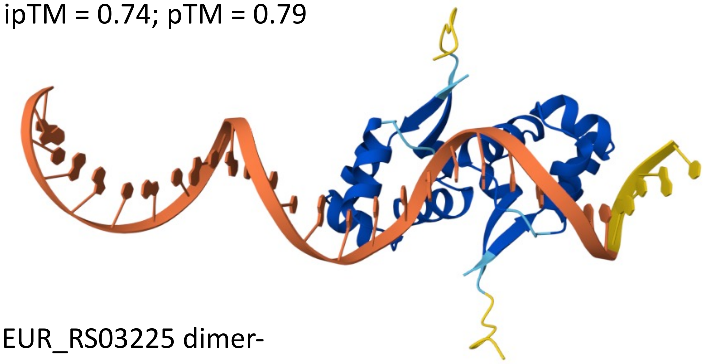

EUR_RS03225 dimer-
*EUR_RS03225*_Mutant:
ipTM = 0.67; pTM = 0.76

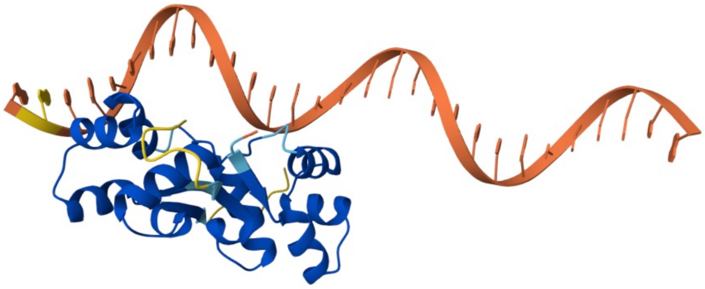

RS03225 dimer-
*EUR_RS01730* _Mutant:
ipTM = 0.69; pTM = 0.77

Figure S15
