## supplementary tables for "Gut microbial structural variation improves disease discrimination and reveals a strain-level regulatory mechanism in rheumatoid arthritis"

Table S1. Clinical and Demographic Characteristics of Patients Across Four Independent Cohorts.

| Characteristic | | Age (years), median (IQR) | Female sex, n (%) | ACPA positivity, n (%) | ESR (median, IQR) | CRP (median, IQR) |
|---|---|---|---|---|---|---|
| Cohort A | RA (n=76) | 60 (53–66) | 61 (80.2%) | 53 (70.0%) | 60.00 (37.60–85.75) | 32.50 (14.50–54.70) |
| | OA (n=19) | 66 (64–71) | 15 (78.9%) | 0 (0%) | 23.00 (15.75–41.50) | 17.45 (4.62–65.75) |
| | HC (n=27) | 56 (50–60) | 19 (70.3%) | 0 (0%) | N/A | N/A |
| Cohort B | RA (n=40) | 52 (42–62)* | 27 (67.5%) | 25 (62.5%) | 44 (28–75)* | 24.8 (8.5–65)* |
| | HC (n=29) | 52 (40–60)* | 20 (69.0%) | 0 (0%) | N/A | N/A |
| Cohort C | RA (n=37) | 56 (52–60)* | 29 (78.4%) | 37 (100%) | 33 (22–45)* | 12.1 (7.5–22.5)* |
| | HC (n=31) | 55 (52–59)* | 24 (77.4%) | 0 (0%) | N/A | N/A |
| Cohort D | RA, Untreated (n=94) | 51 (43-57) | 73 (77.7%) | 62 (66.0%) | 44.00 (23.00-67.00) | 14.15 (6.20-38.62) |
| | RA, Treated (n=41) | 53 (41-57) | 34 (82.9%) | 5 (12.2%) | 20.00 (10.00-29.00) | 3.34 (1.70-6.60) |
| | HC (n=97) | 43 (37-48) | 67 (69.1%) | 0 (0.0%) | N/A | N/A |

Note: * Values are estimated interquartile ranges (IQR) derived from the reported median and range. The estimation follows the formula: IQR ≈ Range × 0.34.

Abbreviations: RA, rheumatoid arthritis; OA, osteoarthritis; HC, healthy controls; ACPA, anti-citrullinated protein antibodies; ESR, erythrocyte sedimentation rate; CRP, C-reactive protein; IQR, interquartile range; N/A, not available.

Table S2. Statistical comparison of alpha diversity metrics across cohorts.

| Cohort | Groups | Shannon Index (*P*) | Simpson Index (*P*) | Observed Richness (*P*) | Pielou's Evenness (*P*) |
|---|---|---|---|---|---|
| A | 3 | 0.09 | 0.10 | <0.01 | 0.18 |
| B | 2 | 0.87 | 0.95 | 0.54 | 0.88 |
| C | 2 | 0.46 | 0.35 | 0.65 | 0.41 |
| D | 3 | 0.43 | 0.45 | 0.24 | 0.58 |

Note: Data represent the *P*-values derived from statistical comparisons across groups within each cohort (e.g., Kruskal-Wallis test for Cohorts A and D with 3 groups; Wilcoxon rank-sum test for Cohorts B and C with 2 groups). $P < 0.05$ is considered statistically significant.

Abbreviations: P, probability value.

Table S3. Statistical analysis of beta diversity using PERMANOVA and beta dispersion tests.

| Cohort | No. of Groups | PERMANOVA | | | Beta dispersion | |
|---|---|---|---|---|---|---|
| | | $R^2$ | F | *P*-value | F | *P*-value |
| A | 3 | 0.04 | 2.46 | <0.01 | 7.13 | <0.01 |
| B | 2 | 0.02 | 1.17 | 0.31 | 4.77 | 0.03 |
| C | 2 | 0.03 | 2.32 | 0.02 | 2.10 | 0.15 |
| D | 3 | 0.01 | 1.16 | 0.26 | 0.53 | 0.59 |

Note: *P*-values were calculated based on Permutational Multivariate Analysis of Variance (PERMANOVA) using 999 permutations to assess the dissimilarities in microbial community structure between groups. $P < 0.05$ is considered statistically significant.

Abbreviations: $R^2$, coefficient of determination; F, F-statistic by permutation; *P*, probability value.

Table S4. Meta-analysis of differentially abundant bacterial species across metagenomic cohorts.

| Species | Meta log2FC | SE | 95% CI Lower | 95% CI Upper | Z-score | P-value | Tau2 | I2 (%) | H2 | Cochran's Q | P (heterogeneity) | No. of Contributing Cohorts | Shift | Mean log2CPM |
|---|---|---|---|---|---|---|---|---|---|---|---|---|---|---|
| Limosilactobacillus fermentum specI_v2_Cluster1050* | 3.42 | 0.44 | 2.56 | 4.29 | 7.73 | <0.01 | 0.00 | 0.00 | 1.00 | 0.07 | 0.785 | 2 | Up | 5.49 |
| Limosilactobacillus fermentum specI_v2_Cluster4855* | 3.26 | 0.43 | 2.41 | 4.10 | 7.53 | <0.01 | 0.00 | 0.00 | 1.00 | 0.10 | 0.748 | 2 | Up | 5.36 |
| Scardovia wiggsiae | 1.36 | 0.26 | 0.85 | 1.86 | 5.27 | <0.01 | 0.00 | 0.00 | 1.00 | 2.53 | 0.283 | 3 | Up | 2.09 |
| Lachnospiraceae bacterium | 0.89 | 0.17 | 0.56 | 1.22 | 5.27 | <0.01 | 0.00 | 0.00 | 1.00 | 2.00 | 0.572 | 4 | Up | 7.58 |
| Leuconostoc mesenteroides | 1.76 | 0.35 | 1.07 | 2.46 | 4.98 | <0.01 | 0.00 | 0.00 | 1.00 | 0.77 | 0.380 | 2 | Up | 3.01 |
| Ethanoligenens harbinense | -1.22 | 0.25 | -1.71 | -0.73 | -4.87 | <0.01 | 0.00 | 0.00 | 1.00 | 1.37 | 0.713 | 4 | Down | 4.23 |
| Serratia liquefaciens | -1.59 | 0.34 | -2.25 | -0.92 | -4.68 | <0.01 | 0.07 | #### | 1.44 | 1.44 | 0.231 | 2 | Down | 2.15 |
| Leuconostoc citreum | 1.58 | 0.34 | 0.91 | 2.24 | 4.66 | <0.01 | 0.00 | 0.00 | 1.00 | 0.47 | 0.493 | 2 | Up | 3.27 |
| Eggerthella sp. | 0.63 | 0.14 | 0.36 | 0.90 | 4.55 | <0.01 | 0.00 | 0.00 | 1.00 | 2.21 | 0.530 | 4 | Up | 4.78 |
| Lacrimispora saccharolytica | -0.74 | 0.17 | -1.06 | -0.41 | -4.42 | <0.01 | 0.01 | #### | 1.13 | 4.27 | 0.234 | 4 | Down | 8.91 |
| Staphylococcus aureus | 1.22 | 0.29 | 0.65 | 1.80 | 4.17 | <0.01 | 0.19 | #### | 2.60 | 7.36 | 0.061 | 4 | Up | 4.84 |
| Streptococcus intermedius | 1.37 | 0.33 | 0.72 | 2.03 | 4.12 | <0.01 | 0.22 | #### | 2.18 | 6.83 | 0.078 | 4 | Up | 2.55 |
| Selenomonas sp. | -1.86 | 0.46 | -2.76 | -0.96 | -4.06 | <0.01 | 0.19 | #### | 1.66 | 1.66 | 0.198 | 2 | Down | 3.50 |

| | | | | | | | | | | | | | | |
|---|---|---|---|---|---|---|---|---|---|---|---|---|---|---|
| Bacteroides fragilis | 1.12 | 0.28 | 0.58 | 1.65 | 4.05 | <0.01 | 0.00 | 0.00 | 1.00 | 0.55 | 0.909 | 4 | Up | 14.64 |
| Xanthomonas euvesicatoria | 2.17 | 0.54 | 1.11 | 3.23 | 4.02 | <0.01 | 0.00 | 0.00 | 1.00 | 0.35 | 0.553 | 2 | Up | 3.07 |
| Eubacterium callanderi | 0.59 | 0.15 | 0.29 | 0.88 | 3.91 | <0.01 | 0.00 | 0.01 | 1.00 | 3.16 | 0.367 | 4 | Up | 6.85 |
| Thomasclavelia spiroformis | 1.74 | 0.47 | 0.82 | 2.66 | 3.70 | <0.01 | 0.50 | #### | 2.76 | 9.35 | 0.025 | 4 | Up | 3.76 |
| Streptococcus agalactiae | 1.05 | 0.29 | 0.49 | 1.62 | 3.65 | <0.01 | 0.00 | 0.00 | 1.00 | 0.16 | 0.689 | 2 | Up | 1.25 |
| Actinomyces viscosus | 1.47 | 0.41 | 0.66 | 2.28 | 3.55 | <0.01 | 0.40 | #### | 3.31 | 7.97 | 0.047 | 4 | Up | 1.97 |
| Actinomyces sp. | 0.38 | 0.11 | 0.16 | 0.60 | 3.43 | <0.01 | 0.00 | 0.00 | 1.00 | 0.93 | 0.819 | 4 | Up | 4.70 |
| Pseudomonas syringae | 2.10 | 0.62 | 0.90 | 3.31 | 3.41 | <0.01 | 0.12 | #### | 1.19 | 1.19 | 0.275 | 2 | Up | 2.97 |
| Streptococcus gordonii | 1.38 | 0.41 | 0.57 | 2.18 | 3.33 | <0.01 | 0.38 | #### | 2.79 | 7.54 | 0.056 | 4 | Up | 4.61 |
| Anaerostipes caccae | -1.28 | 0.39 | -2.05 | -0.52 | -3.29 | <0.01 | 0.00 | 0.00 | 1.00 | 0.02 | 0.894 | 2 | Down | 3.18 |
| Butyricicoccus pullicaecorum | 1.05 | 0.32 | 0.42 | 1.67 | 3.29 | <0.01 | 0.21 | #### | 2.45 | 7.55 | 0.056 | 4 | Up | 3.44 |
| Phocaeicola dorei | -0.72 | 0.24 | -1.18 | -0.26 | -3.04 | <0.01 | 0.06 | #### | 1.37 | 3.43 | 0.330 | 4 | Down | 16.51 |
| Streptococcus vestibularis | 1.29 | 0.44 | 0.43 | 2.16 | 2.94 | <0.01 | 0.42 | #### | 2.55 | 8.19 | 0.042 | 4 | Up | 5.15 |
| Brachyspira pilosicoli | -1.37 | 0.47 | -2.30 | -0.44 | -2.89 | <0.01 | 0.53 | #### | 3.12 | 10.21 | 0.017 | 4 | Down | 4.34 |
| Parascardovia denticolens | 2.02 | 0.71 | 0.63 | 3.41 | 2.86 | <0.01 | 0.91 | #### | 3.03 | 6.01 | 0.050 | 3 | Up | 2.39 |
| [Ruminococcus] lactaris | 1.00 | 0.35 | 0.31 | 1.70 | 2.83 | <0.01 | 0.00 | 0.00 | 1.00 | 0.00 | 0.966 | 2 | Up | 2.62 |
| Megasphaera elsdenii | 1.92 | 0.70 | 0.54 | 3.30 | 2.73 | <0.01 | 0.95 | #### | 2.13 | 6.43 | 0.092 | 4 | Up | 8.55 |

| | | | | | | | | | | | | | | |
|---|---|---|---|---|---|---|---|---|---|---|---|---|---|---|
| Butyrivibrio fibrisolvens | -0.60 | 0.22 | -1.04 | -0.17 | -2.73 | <0.01 | 0.02 | #### | 1.12 | 1.91 | 0.385 | 3 | Down | 1.90 |
| Streptococcus oralis | 0.98 | 0.37 | 0.26 | 1.70 | 2.68 | <0.01 | 0.17 | #### | 1.75 | 3.38 | 0.184 | 3 | Up | 1.49 |
| Alkaliphilus oremlandii | -0.84 | 0.31 | -1.45 | -0.22 | -2.68 | <0.01 | 0.08 | #### | 1.47 | 1.47 | 0.226 | 2 | Down | 0.73 |
| Salmonella bongori | 0.73 | 0.27 | 0.19 | 1.26 | 2.67 | <0.01 | 0.00 | 0.00 | 1.00 | 2.17 | 0.538 | 4 | Up | 4.89 |
| Ligilactobacillus ruminis | -1.21 | 0.47 | -2.13 | -0.29 | -2.57 | 0.01 | 0.45 | #### | 2.29 | 7.23 | 0.065 | 4 | Down | 5.93 |
| Dorea longicatena | 0.62 | 0.25 | 0.13 | 1.10 | 2.47 | 0.01 | 0.08 | #### | 1.50 | 4.41 | 0.220 | 4 | Up | 7.04 |
| Faecalibacterium prausnitzii | -0.39 | 0.16 | -0.70 | -0.08 | -2.44 | 0.01 | 0.01 | 8.78 | 1.10 | 3.00 | 0.392 | 4 | Down | 8.60 |
| Streptococcus sp. specI_v2_Cluster1399** | 1.56 | 0.65 | 0.28 | 2.84 | 2.40 | 0.02 | 0.85 | #### | 4.52 | 7.31 | 0.026 | 3 | Up | 3.52 |
| Gallibacterium anatis | 1.82 | 0.77 | 0.31 | 3.33 | 2.36 | 0.02 | 0.82 | #### | 3.07 | 3.07 | 0.080 | 2 | Up | 2.50 |
| Streptococcus sanguinis | 0.87 | 0.37 | 0.14 | 1.60 | 2.34 | 0.02 | 0.32 | #### | 2.95 | 8.16 | 0.043 | 4 | Up | 4.25 |
| Pseudoselenomonas ruminantium | -2.37 | 1.02 | -4.37 | -0.37 | -2.32 | 0.02 | 1.43 | #### | 4.39 | 4.39 | 0.036 | 2 | Down | 4.91 |
| Selenomonas sputigena | -2.08 | 0.90 | -3.85 | -0.31 | -2.31 | 0.02 | 1.11 | #### | 4.20 | 4.20 | 0.040 | 2 | Down | 3.67 |
| Corynebacterium jeikeium | 1.21 | 0.54 | 0.16 | 2.27 | 2.26 | 0.02 | 0.54 | #### | 3.11 | 6.88 | 0.032 | 3 | Up | 2.96 |
| Lachnoclostridium phytofermentans | -0.36 | 0.16 | -0.68 | -0.05 | -2.25 | 0.02 | 0.00 | 0.00 | 1.00 | 0.35 | 0.951 | 4 | Down | 3.71 |
| Desulfurispirillum indicum | 2.06 | 0.94 | 0.23 | 3.89 | 2.20 | 0.03 | 1.06 | #### | 2.61 | 2.61 | 0.106 | 2 | Up | 2.57 |
| Roseburia hominis | -0.53 | 0.25 | -1.01 | -0.04 | -2.14 | 0.03 | 0.14 | #### | 2.77 | 9.44 | 0.024 | 4 | Down | 13.24 |

| | | | | | | | | | | | | | | |
|---|---|---|---|---|---|---|---|---|---|---|---|---|---|---|
| Streptococcus sp. specI_v2_Cluster5125** | 0.76 | 0.36 | 0.06 | 1.47 | 2.13 | 0.03 | 0.27 | #### | 2.45 | 7.31 | 0.063 | 4 | Up | 2.41 |
| Allobaculum stercoricanis | -0.55 | 0.26 | -1.07 | -0.04 | -2.10 | 0.04 | 0.16 | #### | 2.83 | 9.61 | 0.022 | 4 | Down | 5.57 |
| Ruminococcus callidus | -0.86 | 0.44 | -1.72 | -0.01 | -1.98 | 0.05 | 0.29 | #### | 1.66 | 4.95 | 0.175 | 4 | Down | 5.81 |

Note: Meta-analysis was performed using random-effects models with restricted maximum likelihood (REML) estimation across metagenomic cohorts. Positive Meta log2FC values indicate enrichment in rheumatoid arthritis samples, whereas negative Meta log2FC values indicate depletion.

* Taxonomic annotations were derived from the proGenomes database. These entries share the same NCBI species name (Limosilactobacillus fermentum) but represent distinct specI species clusters (specI_v2_Cluster1050 and specI_v2_Cluster4855, respectively).

** Entries correspond to distinct specI species clusters (specI_v2_Cluster1399 and specI_v2_Cluster5125, respectively) within the Streptococcus genus in the proGenomes database.

Abbreviations: log2FC, log2 fold change of relative abundance; SE, standard error of the pooled effect size; CI, confidence interval; Z-score, Z-statistic; P-value, probability value; Tau2, estimated between-cohort variance; I2, heterogeneity statistic; H2, heterogeneity index; Q, Cochran's Q statistic; P (heterogeneity), P value for heterogeneity test; No. of Contributing Cohorts, number of cohorts showing consistent direction of effect; CPM, counts per million.

Table S5. Meta-analysis of variable structural variants across metagenomic cohorts.

| SV ID | Meta SMD | SE | 95% CI Lower | 95% CI Upper | P-value | P (heterogeneity) | No. of Contributing Cohorts |
|---|---|---|---|---|---|---|---|
| Bacteroides uniformis: 3885_3889 | -0.30 | 0.13 | -0.56 | -0.05 | 0.02 | 0.79 | 4 |
| Bacteroides uniformis: 3889_3890 | -0.29 | 0.13 | -0.55 | -0.03 | 0.03 | 0.86 | 4 |
| Bacteroides uniformis: 3884_3885 | -0.26 | 0.13 | -0.52 | 0.00 | 0.05 | 0.88 | 4 |
| Bacteroides uniformis: 3880_3883 | -0.34 | 0.13 | -0.60 | -0.09 | 0.01 | 0.75 | 4 |
| Oscillibacter sp.: 1889_1891 | -0.48 | 0.19 | -0.86 | -0.10 | 0.01 | 0.96 | 4 |
| Oscillibacter sp.: 1721_1726;1728_1731;2108_2111;2111_2113 | -0.46 | 0.19 | -0.84 | -0.07 | 0.02 | 0.56 | 4 |
| Oscillibacter sp.: 1731_1732 | -0.48 | 0.22 | -0.92 | -0.04 | 0.03 | 0.33 | 4 |
| Oscillibacter sp.: 1726_1728 | -0.51 | 0.25 | -0.99 | -0.03 | 0.04 | 0.25 | 4 |
| Oscillibacter sp.: 1784_1785 | -0.52 | 0.19 | -0.90 | -0.14 | 0.01 | 0.93 | 4 |
| Segatella copri: 77_78 | -0.58 | 0.28 | -1.13 | -0.03 | 0.04 | 0.57 | 4 |
| Faecalibacterium prausnitzii: 2481_2482 | 0.29 | 0.15 | 0.00 | 0.58 | 0.05 | 0.50 | 4 |
| Faecalibacterium prausnitzii: 1382_1385;1385_1387 | -0.37 | 0.15 | -0.66 | -0.08 | 0.01 | 0.99 | 4 |

| | | | | | | | |
|---|---|---|---|---|---|---|---|
| Ruminococcus sp.: 768_769 | -0.94 | 0.43 | -1.78 | -0.10 | 0.03 | 0.53 | 2 |
| Parabacteroides merdae: 2404_2410;2410_2422;2422_2423 | -0.35 | 0.17 | -0.69 | -0.01 | 0.04 | 0.61 | 4 |
| Parabacteroides merdae: 782_783;2923_2924 | -0.45 | 0.17 | -0.79 | -0.11 | 0.01 | 0.66 | 4 |
| Roseburia intestinalis: 1845_1851 | 0.45 | 0.20 | 0.05 | 0.84 | 0.03 | 0.51 | 4 |
| Anaerostipes hadrus: 130_132 | 0.63 | 0.32 | 0.01 | 1.26 | 0.05 | 0.46 | 2 |
| Anaerostipes hadrus: 2814_2816 | 0.63 | 0.32 | 0.01 | 1.26 | 0.05 | 0.92 | 2 |
| Ruminococcus callidus: 2759_2760 | -0.72 | 0.32 | -1.35 | -0.10 | 0.02 | 0.97 | 4 |
| Ruminococcus callidus: 2758_2759;2760_2767 | -0.65 | 0.32 | -1.28 | -0.03 | 0.04 | 0.93 | 4 |
| Ruminococcus callidus: 2440_2442;2447_2450;2450_2451 | -0.70 | 0.32 | -1.33 | -0.07 | 0.03 | 0.83 | 4 |
| Ruminococcus bromii: 97_103 | 0.59 | 0.28 | 0.04 | 1.14 | 0.04 | 0.07 | 4 |
| Phocaeicola massiliensis: 4194_4195;4195_4196;4196_4197;4197_4198;4198_4199;4199_4200;4200_4201;4201_4202 | -0.57 | 0.24 | -1.04 | -0.10 | 0.02 | 0.91 | 4 |
| Bacteroides fragilis: 4781_4788 | -0.79 | 0.31 | -1.40 | -0.18 | 0.01 | 0.76 | 4 |

| | | | | | | | |
|---|---|---|---|---|---|---|---|
| Alistipes shahii: 2252_2255;2255_2257;2257_2259;2259_2262;2262_2264;2265_2266 | -0.38 | 0.18 | -0.73 | -0.03 | 0.03 | 0.48 | 4 |
| [Ruminococcus] lactaris: 1750_1753 | 1.05 | 0.38 | 0.32 | 1.79 | 0.01 | 0.51 | 3 |
| Parabacteroides distasonis: 4288_4291;4291_4293 | 0.36 | 0.16 | 0.04 | 0.67 | 0.03 | 0.32 | 4 |
| Bacteroides caccae: 1351_1363 | -0.41 | 0.18 | -0.77 | -0.05 | 0.03 | 0.97 | 4 |
| Agathobacter rectalis: 585_586 | -0.32 | 0.16 | -0.64 | -0.01 | 0.04 | 0.46 | 4 |
| Agathobacter rectalis: 461_469 | -0.38 | 0.16 | -0.70 | -0.07 | 0.02 | 0.79 | 4 |
| Agathobacter rectalis: 3036_3037 | -0.39 | 0.16 | -0.71 | -0.08 | 0.01 | 0.55 | 4 |
| Agathobacter rectalis: 3102_3105;3105_3106 | -0.34 | 0.16 | -0.65 | -0.03 | 0.03 | 0.85 | 4 |
| Bacteroides xylanisolvens: 3046_3048;3378_3379 | 0.44 | 0.18 | 0.08 | 0.80 | 0.02 | 0.89 | 4 |
| Bacteroides xylanisolvens: 4246_4248;4260_4262;4265_4267;4268_4270;4271_4280 | 0.37 | 0.18 | 0.01 | 0.73 | 0.04 | 0.79 | 4 |

Note: Meta-analysis was performed using random-effects models with restricted maximum likelihood (REML) estimation across metagenomic cohorts. Standardized mean differences (SMD) were calculated for continuous relative abundances of variable structural variants (vSVs). Positive Meta SMD values indicate enrichment in rheumatoid arthritis samples, whereas negative Meta SMD values indicate depletion.

Abbreviations: SV ID, structural variant identifier; Meta SMD, standardized mean difference of relative abundance; SE, standard error of the pooled effect size; CI, confidence interval; P, probability value; P (heterogeneity), P value for heterogeneity test; No. of Contributing Cohorts, number of cohorts showing consistent direction of effect.

Table S6. Meta-analysis of deletion structural variants across metagenomic cohorts.

| SV ID | Meta OR | 95% CI Lower | 95% CI Upper | P-value | P (heterogeneity) | No. of Contributing Cohorts |
|---|---|---|---|---|---|---|
| Blautia wexlerae: 1054_1055;1598_1602 | 0.25 | 0.09 | 0.71 | 0.01 | 0.71 | 2 |
| Phascolarctobacterium sp.: 0_1;228_230;230_244;340_341;891_892 | 2.20 | 1.05 | 4.60 | 0.04 | 0.52 | 2 |
| Oscillibacter sp.: 1356_1357 | 0.30 | 0.11 | 0.81 | 0.02 | 0.48 | 2 |
| Oscillibacter sp.: 557_558;1131_1132 | 2.26 | 1.04 | 4.91 | 0.04 | 0.84 | 2 |
| Anaerostipes hadrus: 1104_1111;1111_1112 | 0.24 | 0.07 | 0.86 | 0.03 | 0.79 | 2 |
| Anaerostipes hadrus: 443_444;903_904 | 0.09 | 0.02 | 0.39 | 0.00 | 0.98 | 2 |
| Anaerostipes hadrus: 895_896;897_898 | 0.21 | 0.05 | 0.90 | 0.04 | 0.63 | 2 |
| Mediterraneibacter gnavus: 637_639;2887_2888 | 0.19 | 0.04 | 0.89 | 0.04 | 0.33 | 2 |
| Bacteroides uniformis: 2461_2495;3120_3121 | 2.01 | 1.18 | 3.43 | 0.01 | 0.97 | 2 |
| Bacteroides uniformis: 2624_2628;4186_4192 | 1.82 | 1.03 | 3.24 | 0.04 | 0.73 | 2 |
| Parabacteroides distasonis: 3054_3055 | 0.35 | 0.16 | 0.76 | 0.01 | 0.49 | 2 |
| [Ruminococcus] lactaris: 320_321;1385_1386 | 0.10 | 0.02 | 0.51 | 0.01 | 0.83 | 2 |

| | | | | | | |
|---|---|---|---|---|---|---|
| Bifidobacterium pseudocatenulatum: 130_131;1081_1082 | 0.27 | 0.09 | 0.81 | 0.02 | 0.59 | 2 |
| Bacteroides xylanisolvens: 5601_5602 | 0.46 | 0.21 | 0.98 | 0.05 | 0.50 | 2 |
| Agathobacter rectalis: 662_663 | 2.49 | 1.26 | 4.92 | 0.01 | 0.83 | 3 |
| Ruminococcus bromii: 290_291;972_973;1878_1879 | 0.34 | 0.15 | 0.80 | 0.01 | 0.53 | 3 |
| Ruminococcus bromii: 37_39 | 3.02 | 1.31 | 6.93 | 0.01 | 0.47 | 2 |
| Alistipes shahii: 2840_2842 | 0.33 | 0.15 | 0.71 | 0.00 | 0.93 | 2 |
| Roseburia intestinalis: 2079_2081 | 3.05 | 1.17 | 7.95 | 0.02 | 0.31 | 2 |
| Roseburia intestinalis: 3179_3180 | 0.24 | 0.07 | 0.86 | 0.03 | 0.14 | 2 |
| Barnesiella intestinihominis: 324_326;326_328;328_330 | 0.17 | 0.05 | 0.57 | 0.00 | 0.73 | 2 |
| Barnesiella intestinihominis: 3324_3326 | 0.30 | 0.11 | 0.87 | 0.03 | 0.71 | 2 |
| Faecalibacterium prausnitzii: 537_538 | 1.89 | 1.01 | 3.54 | 0.05 | 0.30 | 2 |
| Faecalibacterium prausnitzii: 781_783 | 2.35 | 1.27 | 4.36 | 0.01 | 0.98 | 3 |
| Faecalibacterium prausnitzii: 847_849 | 2.47 | 1.08 | 5.66 | 0.03 | 0.86 | 2 |

Note: Meta-analysis was performed using random-effects models with the Mantel-Haenszel (MH) method across metagenomic cohorts. Odds ratio (OR) values were pooled for deletion structural variants (dSVs). OR > 1 indicates increased odds/risk in rheumatoid arthritis samples, whereas OR < 1 indicates decreased odds.
Abbreviations: SV ID, structural variant identifier; Meta OR, pooled odds ratio; 95% CI, 95% confidence interval; P, probability value; P (heterogeneity), P value for heterogeneity test; No. of Contributing Cohorts, number of cohorts showing consistent direction of effect.

Table S7. Performance evaluation and comparison of machine learning models incorporating species and structural variations.

| Model Type | No. of Features | Lambda Value | Test AUC (95% CI) | DeLong's P-value |
|---|---|---|---|---|
| Species-only (λ.1se) | 3 | 0.09 | 0.476 (0.330–0.622) | 0.04 |
| Species+SV (λ.1se) | 52 | 0.02 | 0.621 (0.480–0.761) | |
| Species-only (λ.min) | 20 | 0.03 | 0.449 (0.308–0.590) | <0.01 |
| Species+SV (λ.min) | 58 | 0.01 | 0.603 (0.462–0.745) | |

Note: Machine learning prediction models were constructed using penalized logistic regression (Lasso) on the training dataset (Cohort B+C). Optimal penalty parameters were selected based on the 1-standard-error rule (λ.1se) and minimum cross-validation error (λ.min). Model performance (AUC) and statistical significance of AUC differences (DeLong's test) were independently evaluated on the validation dataset (Cohort D).

Abbreviations: AUC, area under the receiver operating characteristic curve; CI, confidence interval; SV, structural variant; λ.1se, lambda value corresponding to 1 standard error above the minimum cross-validation error; λ.min, lambda value that minimizes mean cross-validated error.

Table S8. Feature importance and directional correlation of key structural variants across training and independent validation cohorts.

| SV ID | Mean \|SHAP\| (Training) | Mean \|SHAP\| (Test) | Spearman r (Training) | P-value (Training) | Spearman r (Test) | P-value (Test) | Effect (Training) | Effect (Test) | Directional Consistency |
|---|---|---|---|---|---|---|---|---|---|
| Alistipes shahii: 2252_2255;2255_2257;2257_2259;2259_2262;2262_2264;2265_2266 | 0.03 | 0.02 | 0.89 | <0.01 | 0.75 | <0.01 | Promote RA (Risk) | Promote RA (Risk) | Consistent |
| Faecalibacterium prausnitzii: 537_538 | 0.01 | 0.02 | -0.70 | <0.01 | -0.81 | <0.01 | Protect against RA (Protective) | Protect against RA (Protective) | Consistent |
| Ruminococcus bromii: 97_103 | 0.01 | 0.01 | -0.89 | <0.01 | -0.84 | <0.01 | Protect against RA (Protective) | Protect against RA (Protective) | Consistent |
| Bacteroides uniformis: 3880_3883 | 0.01 | 0.01 | 0.82 | <0.01 | 0.85 | <0.01 | Promote RA (Risk) | Promote RA (Risk) | Consistent |
| Faecalibacterium prausnitzii: 781_783 | 0.01 | 0.01 | -0.71 | <0.01 | -0.77 | <0.01 | Protect against RA (Protective) | Protect against RA (Protective) | Consistent |
| Bacteroides fragilis: 4781_4788 | 0.01 | 0.00 | 0.91 | <0.01 | 0.69 | <0.01 | Promote RA (Risk) | Promote RA (Risk) | Consistent |
| Agathobacter rectalis: 3036_3037 | 0.01 | 0.00 | 0.81 | <0.01 | 0.72 | <0.01 | Promote RA (Risk) | Promote RA (Risk) | Consistent |
| Agathobacter rectalis: 662_663 | 0.01 | 0.01 | -0.67 | <0.01 | -0.77 | <0.01 | Protect against RA (Protective) | Protect against RA (Protective) | Consistent |
| Oscillibacter sp.: 557_558;1131_1132 | 0.01 | 0.01 | -0.76 | <0.01 | -0.75 | <0.01 | Protect against RA (Protective) | Protect against RA (Protective) | Consistent |
| Agathobacter rectalis: 3102_3105;3105_3106 | 0.00 | 0.00 | 0.67 | <0.01 | 0.72 | <0.01 | Promote RA (Risk) | Promote RA (Risk) | Consistent |
| Bacteroides xylanisolvens: 5601_5602 | 0.00 | 0.00 | 0.68 | <0.01 | 0.66 | <0.01 | Promote RA (Risk) | Promote RA (Risk) | Consistent |
| Parabacteroides merdae: 782_783;2923_2924 | 0.00 | 0.00 | 0.70 | <0.01 | 0.81 | <0.01 | Promote RA (Risk) | Promote RA (Risk) | Consistent |

Note: Key structural variants (SVs) were rigorously filtered based on four consensus criteria: (1) ranking among the top 20 most important features evaluated by mean absolute SHAP values (Mean |SHAP|); (2) demonstrating strong monotonic correlation with rheumatoid arthritis (RA) status (|Spearman r| ⩾ 0.6); (3) achieving strict statistical significance ($P < 0.001$) in both the training dataset (Cohort B+C) and the independent validation dataset (Cohort D); and (4) showing 100% directional consistency between training and validation cohorts.

Abbreviations: SV ID, structural variant identifier; SHAP, SHapley Additive exPlanations; r, Spearman's rank correlation coefficient; P, probability value; RA, rheumatoid arthritis; Training, Cohort B+C; Test, Cohort D.
